# Remodeling of human diurnal adipose tissue transcriptome by the composition of morning and afternoon meals

**DOI:** 10.1101/2025.05.14.25327610

**Authors:** Jorge R. Soliz-Rueda, Katharina Kessler, Karsten Jürchott, Carsten Sticht, Silke Hornemann, Achim Kramer, Andreas F. H. Pfeiffer, Olga Pivovarova-Ramich

## Abstract

The timing of carbohydrate and fat intake modulates the metabolic status, yet the underlying mechanisms in humans remain poorly understood. In this crossover trial, we investigated the effects of two isocaloric 4-week dietary patterns - high carbohydrate in the morning and high fat in the afternoon (HC/HF) and the reverse (HF/HC) - on the subcutaneous adipose tissue (SAT) transcriptome in non-diabetic men. Analysis of SAT samples collected across the day identified 1386 genes exhibiting diurnal oscillations. Both oscillatory and non-oscillatory genes related to lipid and glucose metabolism were modulated by the timing of macronutrient intake, in SAT as well as in peripheral blood monocytes. Notably, expression of inflammatory response genes in SAT was elevated after HC/HF compared to HF/HC. These findings demonstrate that diurnal macronutrient distribution significantly reshapes the SAT transcriptome, underscoring the relevance of chrononutrition as a potential strategy to prevent metabolic dysfunction and systemic inflammation associated with obesity and type 2 diabetes.

## Introduction

Components of circadian and metabolic cycles in mammals are tightly interrelated^1^. The circadian clocks in metabolically active tissues control the expression of genes crucial for energy homeostasis and allow adaptation to the daily light/dark cycles and the resulting changes in food availability^2,3^. Diurnal oscillations in carbohydrate metabolism (gluconeogenesis, glucose uptake, utilization and hormone abundance) are described in animals and humans^4,5^. Similarly, lipid metabolism in adipose tissue (AT), including lipolysis, lipogenesis and endocrine function, is also regulated by the circadian system^6–8^.

Extensive evidence showed that the timing of food intake is an important factor of metabolic regulation^9^. Food intake at times misaligned with natural feeding time resets peripheral clock and disrupts metabolic homeostasis, as has been shown in nocturnal animals fed in the light phase and in shift working humans^10–14^. The timing of the main meal throughout the day influences the risk of obesity and the success of weight loss therapy^9,15^. Many studies showed that late eating is associated with weight gain, dysfunction in energy expenditure, and abnormalities in the circadian rhythms of appetite, stress, and sleep hormones^16^. Human experimental studies demonstrated a better insulin sensitivity, beta cell responsiveness, glucose tolerance, an increased postprandial thermogenesis and different postprandial hormone secretion in response to the morning meal compared to the same meal consumed in the afternoon/evening^17–19^.

These findings raise the hypothesis that composition of the morning and afternoon meals, e.g. carbohydrate and fat intake, might modulate metabolic status in humans. In fact, mice fed a high-carb (HC) food at the beginning of the active period and a high-fat (HF) food at the end showed increased body weight and blood glucose compared to opposite food distribution during the active phase^20^. In humans, long-term studies on metabolic effects in response to carbohydrate and fat consumption at different day time are sparse and controversial^21–23^. Epidemiological studies propose a beneficial effect of a carbohydrate-rich diet at the beginning of the day which is shown to be protective against the development of diabetes and metabolic syndrome^24^. In agreement with this, we have recently shown in a CLOCK trial (NCT02487576) that a diet where fat is mainly eaten in the morning and carbs mainly in the afternoon (compared to the reverse order) worsens glycemic control in people with prediabetes^18^. We have also found that the composition of morning and afternoon meals affects daily profiles of substrate oxidation and other circulating metabolic markers (lipids, adipokines and cytokines) in humans^18,25^, but molecular mechanisms involved in this regulation are poorly understood. In this context, many studies in mice have demonstrated the ability of diet composition to modify the oscillatory transcriptome expression pattern of different peripheral tissues, including white AT^26–28^.

To investigate the impact of diurnal macronutrient distribution on the tissue-specific transcriptome in humans, we here studied how carbohydrate and fat intake during the morning and evening meals affects the daily pattern of AT transcriptome and which signaling pathways are involved in this regulation. The altered genes and signaling pathways were additionally evaluated in peripheral blood mononuclear cells (PBMCs).

## Results

### Study design and subject characteristics

To investigate the effects of morning and afternoon meal composition on AT transcriptome, a crossover study was conducted in overweight non-diabetic men in a sub-cohort of the CLOCK trial (**Figure S1**). We compared the effects of two 4-week isocaloric diets – (1) an HC/HF diet, which consisted of a high-carb breakfast and lunch (65 EN% carbohydrates, 20 EN% fat, 15 EN% protein) and a high-fat snack and dinner (35 EN% carbohydrates, 50 EN% fat, 15 EN% protein) and (2) an HF/HC diet, which consisted of a high-fat breakfast and lunch and a high-carb snack and dinner (**Figure 1A**). On the investigation day, at the end of each intervention period, two standardized equicaloric meals – high-carb (MTT-HC) or high-fat (MTT-HF) – were administered at 09:00 and 15:40 h, respectively, in an order determined by the participants’ preceding diet.

**Figure 1.**
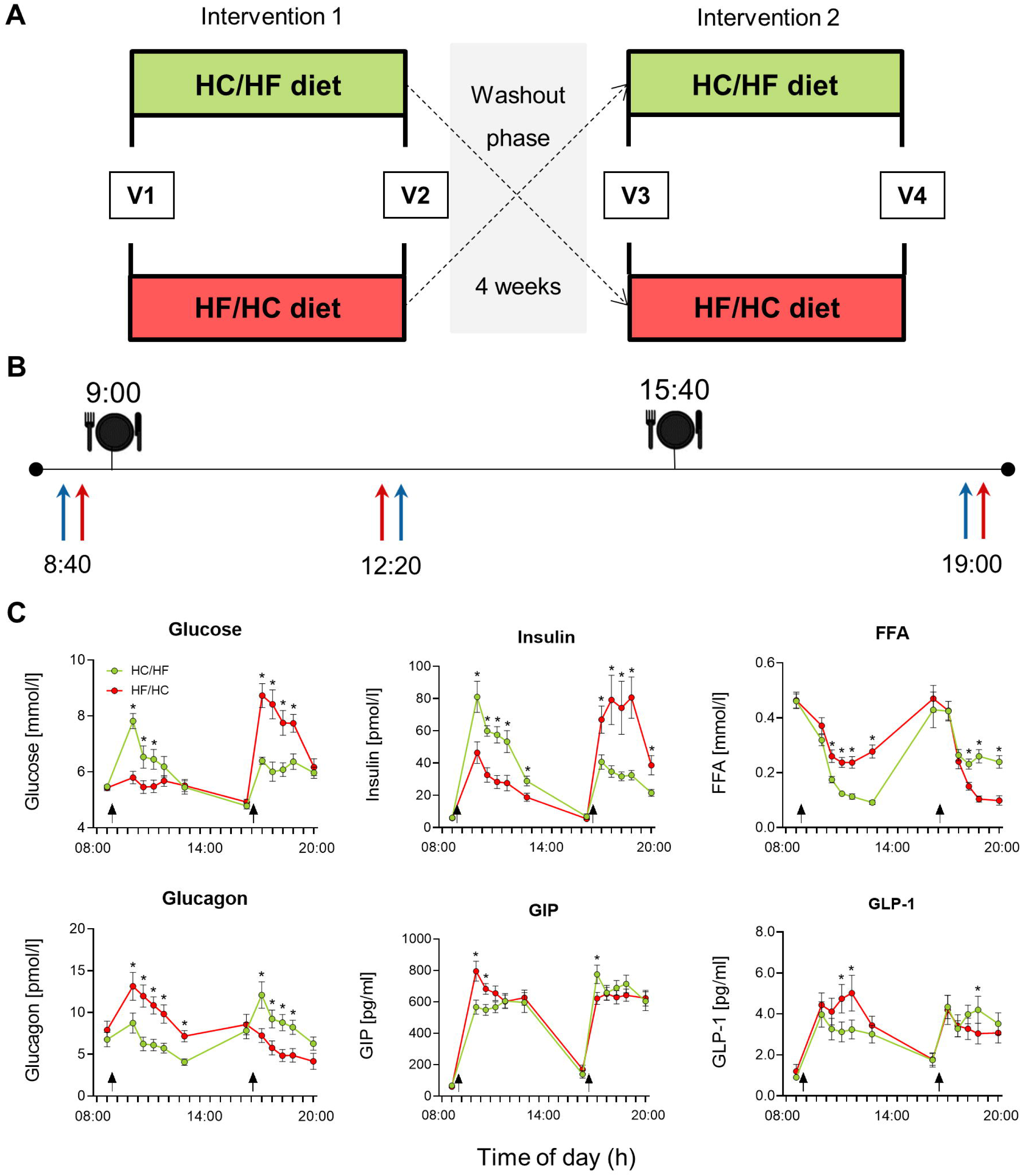
Study design and clinical investigation day. (A) Study design. HC/HF diet, isocaloric high-carbohydrate meals from 6:00 to 13:30 and isocaloric high-fat meals between 16:30 and 22:00; HF/HC diet, reversed order of meal sequence; V, visit. (B) Clinical investigation day. At 09:00 and 15:40 standardized equicaloric test meals – high-fat and high-carbohydrate – were provided in order according to each participant’s previous intervention. Arrows indicate the collection of SAT and PBMC samples. (C) Serum glucose, insulin, free fatty acid (FFA), glucagon, glucose-dependent insulinotropic peptide (GIP) and glucagon-like peptide-1 (GLP-1) levels in serum before and after each test meal (HC/HF diet – green line, HF/HC diet – red line, n=15). Arrows indicate test meals. * p<0.05 indicate significant difference between HC/HF diet vs. HF/HC diet at the same time of the day by paired t-test. Data are expressed as mean ± SEM for each time point.

Subcutaneous AT (SAT) samples were collected at three specified times throughout the day (at 8:40, 12:20 and 19:00 h) (**Figure 1B**) from 15 men (age 44.9 years, BMI 27.5 kg/m^2^). Their anthropometric characteristics, with exception of waist circumference, as well as parameters related to glucose and lipid metabolism, daily calorie intake and sleep time and duration, showed no differences between dietary interventions (**Table S1**). The whole-body insulin sensitivity index HOMA-IR was not affected by the diurnal distribution of macronutrients, whereas rQUICKI (fasting revised quantitative insulin sensitivity check index) (p=0.044) and the adiponectin/leptin ratio throughout the day (p=0.002 by diet*time interaction), widely used as a measure of AT-specific insulin sensitivity^29,30^, were higher after HF/HC diet, indicating better insulin sensitivity (**Table S1**, **Figure S2A**). Dietary macronutrient distribution during both interventions was very close to the prescribed dietary patterns suggesting high adherence to the interventions (**Table S1**). As expected, postprandial glucose and insulin levels were lower, while free fatty acid (FFA), glucagon, glucose-dependent insulinotropic peptide (GIP) and glucagon-like peptide-1 (GLP-1) levels were higher after the MTT-HF compared with MTT-HC (**Figure 1D**).

### Timing of the carb and fat intake reprograms the diurnal pattern of the SAT transcriptome

A total of 25582 genes were detected by microarray analysis of SAT samples. In the two-way repeated measures ANOVA analysis, 3194 (12.5%) genes exhibited a time effect, 3638 (14.2%) showed a diet effect, and 350 (1.4%) genes showed a time*diet interaction (p<0.01) (**Figure 2A, Supplementary Dataset 1**). Genes demonstrating time or time*diet effects were analyzed using a rhythm prediction method^31^ to predict the 24-hour rhythmicity based on the three-time-point data and the *compareRhythms* approach^32^ (**STAR Methods**) to compare gene oscillations between two diets. This approach revealed 1386 genes with a significant cosine rhythm (p<0.05, amplitude>0.2) and assessed their acrophase and amplitude. 995 (71.8%) genes showed the same rhythmicity after both HC/HF and HF/HC diets, and 391 (28.2%) genes were differentially rhythmic (diffR) between two diets. Specifically, the diffR genes were 114 (8.2%) genes that oscillated exclusively after the HC/HF diet, 192 (13.9%) genes that showed diurnal oscillations only after the HF/HC diet, and 85 (6.1%) genes that changed their diurnal oscillation depending on diet (**Figure 2B, Supplementary Dataset 2**).

**Figure 2.**
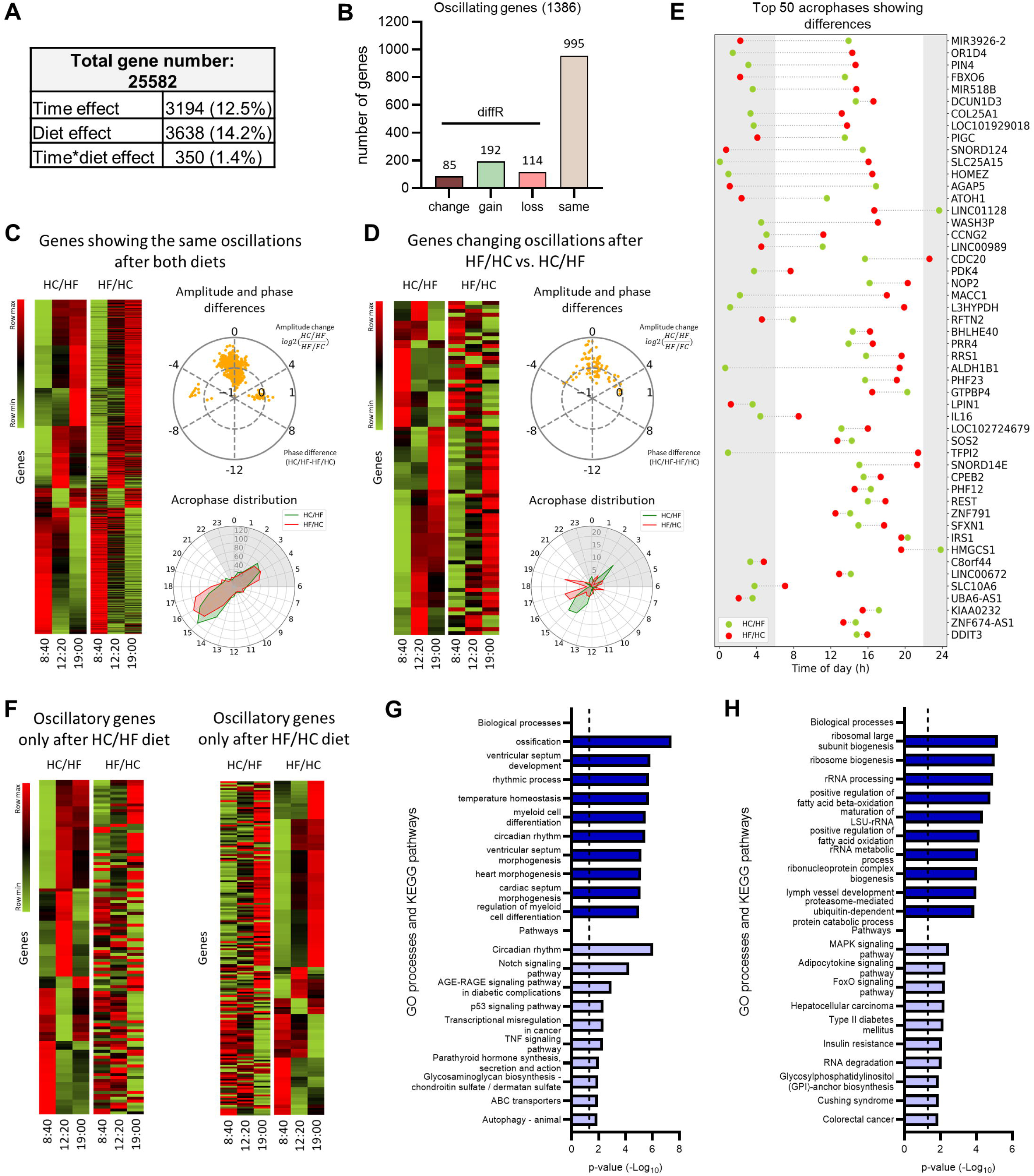
Timing of the carb and fat intake reprograms the diurnal transcriptome in SAT. (A) Number of genes showing a diet effect, time effect or time*diet interaction in a two-way repeated measures ANOVA (p<0.01, without correction for multiple testing). (B) Number of genes oscillating after both diets, exclusively after the HC/HF diet (green), or after the HF/HC diet (red) assessed by a three time point rhythm prediction method (p<0.05, amplitude>0.2). (C) Heatmap representation of genes showing the same oscillations after the both diets and genes changing oscillations after the HF/HC vs HC/HF diet. Amplitude and phase differences and Acrophase distribution for same oscillations upon the both diets and genes changing between two conditions. (D) Top 50 peak times of genes showed a phase difference between two diets by cosinor method (p<0.05, without correction for multiple testing). (E) Heatmap representation of genes oscillating only after the HC/HF and HF/HC diets. (F-G) GO processes and KEGG analysis of oscillating genes (F) after both diets, (F) genes changing after HF/HC vs HC/HF diet as identified by WebGestalt. The full pathway list including gene names is shown in **Table S2**. (A-I) n=15.

The heatmap revealed an overall similar rhythmic pattern of genes exhibiting the same oscillations after both diets (**Figure 2C**). Most of these genes peaked in the morning between 3:00 and 5:00 (18.7% and 17% for HC/HF and HF/HC diet, respectively) and in the afternoon between 14:00 and 17:00 (33.8% and 29.9% for HC/HF and HF/HC diet, respectively), and the number of the afternoon-peaked genes was approximately two-fold higher as the number of morning-peaked genes (**Figure 2C**). When the expression profile at each time points of oscillating genes were analysed separately, the hierarchical clustering analysis of the top 50 oscillatory genes showed a robust separation by diet at 19:00, while this clustering by diet was not observed at 8:40 or 12:20 (**Figure S3**). Functional annotations of genes exhibiting the same oscillation after both diets included circadian rhythm, notch signaling, AGE-RAGE pathway, p53 pathway, as well as TNF signalling. These genes are also involved in different biological processes such as ossification, ventricular septum development, rhythmic process, temperature homeostasis as well as myeloid cell differentiation (**Figure 2G, Table S2, Supplementary Dataset 3**).

A change in the expression pattern was observed in the heatmap for genes that showed a changed oscillation (**Figure 2D**). Regarding the phase distribution of these genes, while an approximately equal number of genes peaked in the morning and in the afternoon upon the HC/HF diet, more oscillating genes exhibited an afternoon peak than morning peak upon the HF/HC diet (**Figure 2D**). 62 genes showed significant differences in peak time between two diets (**Figure 2E, Table S3**), with 40 genes showing a phase advance, and 22 genes showing a phase delay after the HF/HC diet compared to the HC/HF diet. Notably, this shift cannot be explained by the usual meal schedule of subjects, time of test meals during the investigation day or by the difference of the gastric emptying rate between high-carb and high-fat meals (**Figure S2B**). In addition, 14 genes showed a diet-dependent change in the amplitude of their oscillation (**Table S3**), including key genes involved in the insulin signalling and glucose metabolism *IRS1, IRS2, PCK1* (**Figure 4B**) as described in detail below. For diet-specific oscillating genes, the heatmaps showed a rhythmic pattern after one diet, which was lost after the opposite diet (**Figure 2E**). DiffR genes (i.e. genes showing the change, gain or loss of rhythmicity) between two diets were involved in MAPK signaling pathways, adipocytokine signaling, FoxO signaling, type II diabetes mellitus, insulin resistance, as well as glycosylphosphatidylinositol-anchor biosynthesis, and in different biological processes such as ribosomal large subunit biogenesis, ribosome biogenesis, rRNA processing, positive regulation of fatty acid oxidation and beta-oxidation as well as rRNA metabolic process (**Figure 2G, Table S2, Supplementary Dataset 3**).

### Timing of the carb and fat intake does not affect SAT clock and central clock markers

As expected, we found diurnal oscillations of most core clock genes in SAT **(Figure 3A).** Clock genes *BMAL1, CLOCK, PER1, PER2, PER3, CRY1, CRY2, NR1D1, NR1D2, DBP,* and *TEF* showed 24-hour rhythmicity after both diets (**Figure 3A, Table S4)**. In agreement with previously published data^33^, *NR1D1, NR1D2, PER1, PER3, DBP,* and *TEF* genes demonstrated the highest expression in the morning, whereas *BMAL1* and *CLOCK* genes peaked in the evening (**Figure 3A**). The *CRY1* gene had peaks in the early afternoon and *PER2* and *CRY2* genes at noon. Overall, no diet-dependent differences were found in the expression pattern and peak shift of the clock genes (**Table S4**). However, *NR1D1*, *PER2* and *TEF* (p=0.008, p=0.026 and p=0.033, respectively) showed a decrease in the amplitude of their diurnal oscillation upon the HC/HF diet compared to the HF/HC diet (**Figure 3A**, **Table S4**).

**Figure 3.**
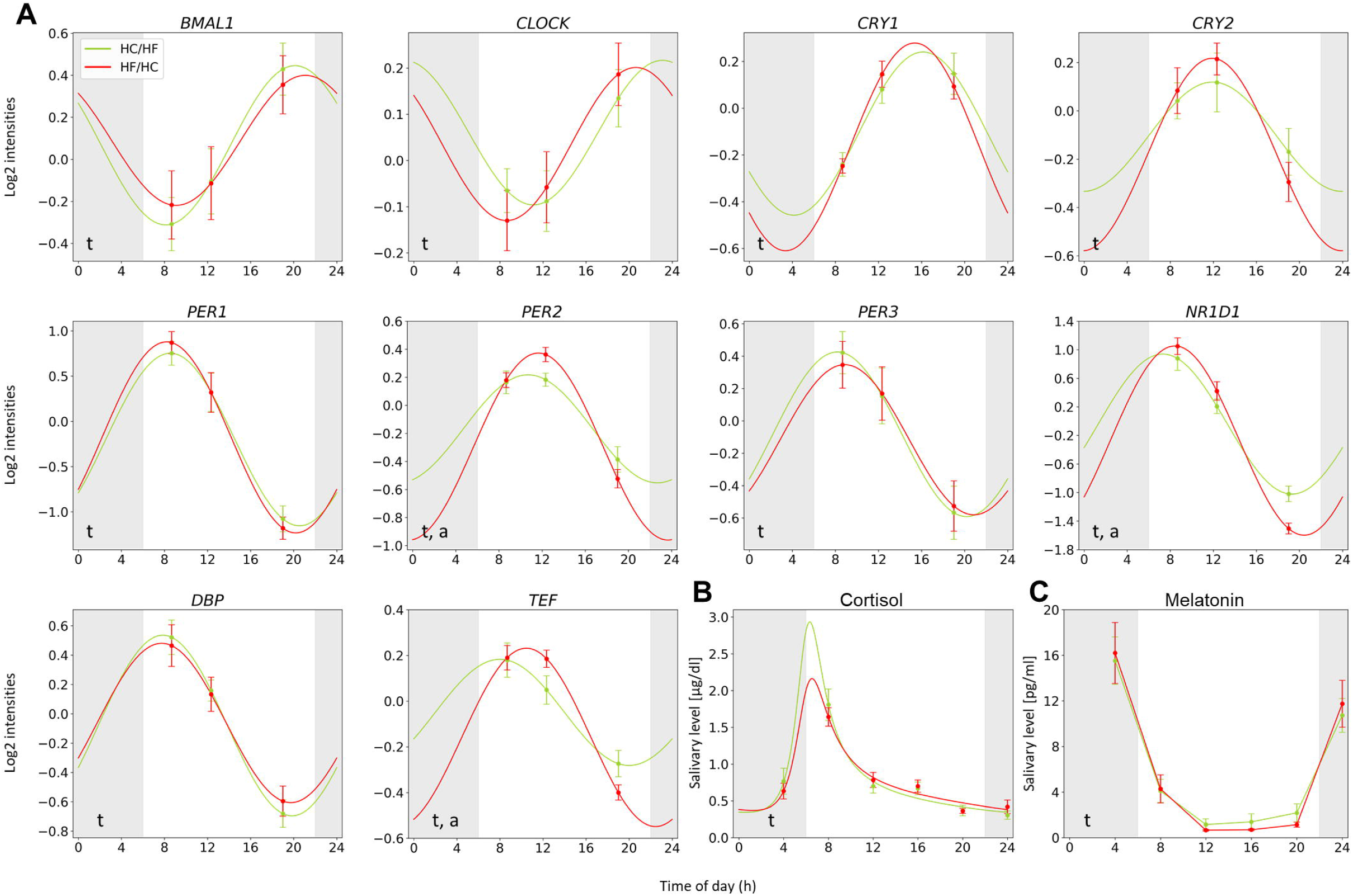
Timing of the carb and fat intake does not affect SAT clock and central clock markers. (A) Diurnal oscillations of clock genes in SAT after HC/HF (green) and HF/HC (red) diets. (B) Salivary cortisol rhythmicity after HC/HF (green) and HF/HC (red) diets. (C) Salivary melatonin levels throughout the day after HC/HF (green) and HF/HC (red) diets. (A-C) n=15. Data are expressed as mean ± SEM for each time point. The oscillation fit was performed using a sinus model (cosinor method). Significant oscillation was considered when amplitude ≥ 0.2 and p<0.05. Significant changes of the oscillation acrophase (p) and/or amplitude (a) are shown. For cortisol, a nadir (time of the curve minimum) was calculated. The full data analysis is shown in **Table S4**. HC, high-carb; HF, high-fat.

To determine an influence of the diurnal carb and fat distribution on the central clock in humans, we measured salivary cortisol and melatonin concentrations over 24 h after the HC/HF and HF/HC diets. As expected, both hormones displayed diurnal variation with a cortisol peak in the morning, approximately at 7:00, and melatonin peak between 0.00 and 4.00 (**Figure 3B-C**). Analysis of cortisol rhythmicity using an extended sinus model^31^ revealed no dietary-dependent differences of its oscillation parameters (**Figure 3B, Table S4**). For salivary melatonin, we were not able to find a fitting model^34^ providing a reasonable accuracy of diurnal rhythm analysis (**Figure 3C**).

### Timing of the carb and fat intake influences oscillating and non-oscillating genes of glucose and lipid metabolism in SAT

We further focused on dietary effects on SAT pathways involved in glucose and lipid metabolism and inflammation playing a key role in the pathogenesis of obesity and type 2 diabetes^35^. For this, a gene set enrichment analysis (GSEA) was performed (**Figure 4A**, **Supplementary Dataset 4**). Most of significantly affected metabolic pathways showed a positive normalized enrichment score (NES), indicating an upregulation of these pathways after the HF/HC diet compared to HC/HF diet (**Figure 4A, Supplementary Dataset 4**). Thus, upon the HF/HC diet, pyruvate metabolism, citrate cycle, oxidative phosphorylation, valine, leucine and isoleucine degradation and fatty acid elongation or degradation, among other metabolic pathways, were affected at all time points (**Figure 4A**).

**Figure 4.**
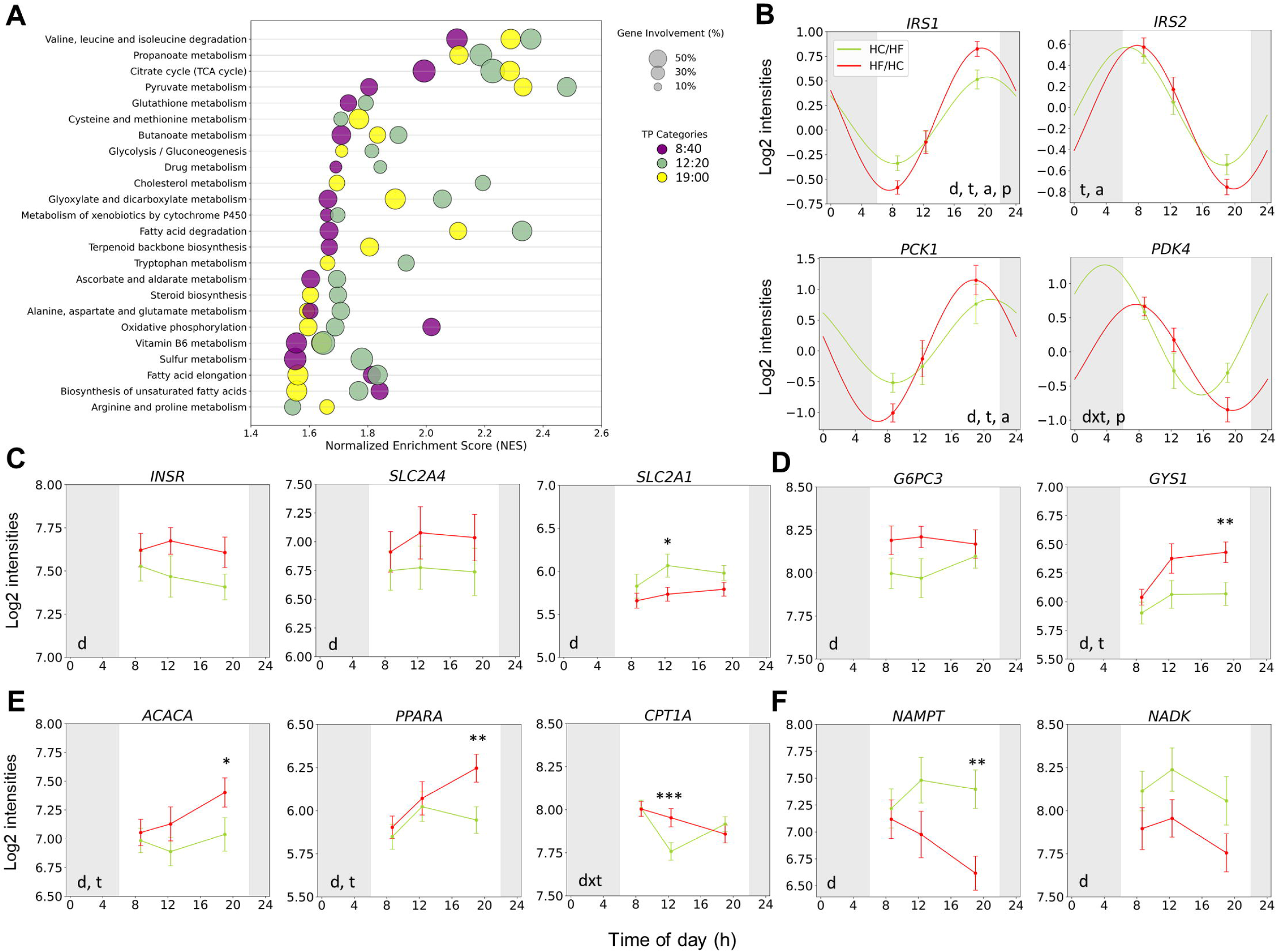
Timing of the carb and fat intake influences oscillating and non-oscillating genes of glucose and lipid metabolism in SAT. (A) Bubble plot representing metabolic pathways changed at 8:40 (purple), 12:20 (green), and 19:00 (yellow) after the HC/HF and HF/HC diets as revealed by GSEA. (B) Diurnal oscillations of glucose and lipid metabolism genes after the HC/HF (green) and HF/HC (red) diets. Diurnal expression patterns are presented in **Figure S4C**. (C) Diurnal expression patterns of non-oscillating insulin signalling genes after the HC/HF (green) and HF/HC (red) diets. (D) Diurnal expression patterns of non-oscillating gluconeogenesis and glycogen metabolism genes after the HC/HF (green) and HF/HC (red) diets. *GSY1* gene showed diurnal oscillation only upon HF/HC diet (**Figure S4E**; **Supplementary Dataset 2**). (E) Diurnal expression patterns of non-oscillating lipogenesis and lipid oxidation genes after the HC/HF (green) and HF/HC (red) diets. (F) Diurnal expression patterns of non-oscillating genes involved in nicotinamide metabolism after the HC/HF (green) and HF/HC (red) diets. (A-G) n=15. Data are expressed as mean ± SEM for each time point. For oscillation genes, the oscillation fit was performed using a sinus model (cosinor method). Significant oscillation was considered when amplitude ≥ 0.2 and p<0.05. Significant changes of the oscillation acrophase (p) and/or amplitude (a) are shown. The full data analysis is shown in **Table S3**. For non-oscillating genes, *p*-values were calculated by two-way repeated-measures ANOVA (d, by diet; t, by time). Genes which did not show diurnal oscillations on one or both diets were presented, the full oscillation analysis is presented in **Supplementary Dataset 2**. * *p*<0.05, ** *p*<0.01 indicate significant difference between HC/HF diet vs. HF/HC diet at the same time of the day by paired t-test. HC, high carbohydrate; HF, high fat; GSEA, gene set enrichment analysis.

As mentioned above, a range of genes representing key nodes in both glucose and lipid metabolism demonstrated diurnal oscillations (**Table S2**, **Supplementary Dataset 2**). For instance, genes involved in lipogenesis, such as sterol regulatory element binding transcription factor 1 (*SREBF1*), diacylglycerol acyltransferase 2 (*DGAT2*) and ATP citrate lyase (*ACLY*), showed diurnal oscillations after both diets with an acrophase occurring during the evening (**Figure S4A-S4C**, **Supplementary Dataset 2**), while genes related to lipid mobilization, including pyruvate dehydrogenase kinase 4 (*PDK4)*, growth arrest and DNA damage inducible beta (*GADD45B*) and sestrin 1 (*SESN1*), showed a peak in the morning (**Figure S4B**, **Supplementary Dataset 2**). The oscillation of some of these genes was affected by diurnal carbohydrate and fat distribution. For instance, genes involved in the glucose metabolism such as insulin receptor substrate 1 (*IRS1)*, insulin receptor substrate 2 (*IRS2)* (p=0.002 and p=0.040, respectively), and the gluconeogenesis gene phosphoenolpyruvate carboxykinase 1 (*PCK1)* (p=0.006) showed changes in the oscillation amplitudes (**Figure 4B-S4D**, **Table S3**). In addition, *IRS1* also showed changes in their acrophases (p=0.008) and the *PDK4* gene, which plays a crucial role in metabolic flexibility, insulin sensitivity and lipid storage, exhibited a 4h phase shift between HC/HF (3:44) and HF/HC (7:37; p<0.001) (**Figure 4B**, **Table S3**).

Diurnal macronutrient distribution also affected the daily expression pattern of non-oscillating genes related to glucose (**Figure 4C-D**) or lipid (**Figure 4E**) metabolism. For instance, insulin signaling pathway genes such as insulin receptor (*INSR*) (p=0.006, here and further in this paragraph: p-values for the diet effect are presented), insulin-responsive glucose transporter type 4 (*SLC2A4/GLUT4*) (p=0.009) showed an increased expression in HF/HC diet, while *SLC2A1/GLUT1* was increased under HC/HF diet (p=0.002) (**Figure 4C**). Further, key genes of glycolysis and glucose utilization pathways (*HK1*, *PKM*, *ALDOA*, *GPI*, *PDHA1*) were affected by timing of the carb and fat intake (**Figure S4E**). Additionally, the gluconeogenesis gene glucose-6-phosphatase catalytic subunit 3 (*G6PC3*) (p=0.008) and glycogen synthase 1 (*GYS1*) (p<0.001) showed an overall expression increase throughout the day upon HF/HC diet by two-way ANOVA (**Figure 4D**). Interestingly, genes related to mitochondrial electron transport and oxidative phosphorylation (*UQCRC2*, *NDUFB5*) were also affected by daily eating patterns, showing an increased expression after HF/HC diet (**Figure S4F**).

Non-oscillating genes involved in lipogenesis, such as acetyl-CoA carboxylase alpha (*ACACA*) (p=0.003), and acetyl-CoA acetyltransferase 1 (*ACAT1*) (p=0.010), and genes involved in fatty acid oxidation, such as peroxisome proliferator-activated receptor alpha (*PPARA*) (p=0.006), acyl-CoA oxidase 2 (*ACOX2*) (p=0.009) and carnitine palmitoyltransferase 1A (*CPT1A*) (p=0.001 by diet*time effect) exhibited similar changes, with an overall increase in gene expression upon the HF/HC diet by two-way ANOVA (**Figure 4E**, **Figure S4G**).

Other energy metabolism genes affected by daily dietary patterns closely related to the circadian core clock^36^ were nicotinamide phosphoribosyltransferase (*NAMPT*) and NAD kinase (*NADK*) genes, whose expression increased throughout the day under HC/HF diet (p=0.001 and p=0.003, respectively) by two-way ANOVA (**Figure 4E**).

### Timing of the carb and fat intake markedly changes inflammatory signaling in SAT

Surprisingly, a large number of inflammatory genes in SAT were also affected by diurnal carb and fat distribution. GSEA revealed affected pathways implicated in leucocyte activation including T cell, B cell and macrophage activation, adhesion and migration, cytotoxicity, phagocytosis, and cytokine production as well as MAPK and NF-kappa B signalling pathways (**Figure 5A**, **Supplementary Dataset 4**). All the pathways involved in the inflammatory response showed a negative NES (< -1.5) at all time points, indicating an up-regulation of the of tissue inflammation upon the HC/HF diet compared to HF/HC (**Figure 5B**, **Supplementary Dataset 4**). Indeed, the diurnal macronutrient distribution affected the expression of inflammatory and chemotactic signalling components, such as the cytokine genes chemokine C-C ligand 5 (*CCL5*) (p<0.001), interleukin-1 beta (*IL1B*) (p<0.001) (**Figure 5B**) and interleukin-16 (*IL16*) (p<0.001) (**Figure S5A**), demonstrating an increased level after the HC/HF diet by two-way ANOVA (**Supplementary Dataset 2**). Interestingly, IL16 also exhibited a diet-dependent shift in acrophase (**Figure S5A, Table S3**). In addition, components involved in the inflammasome development, e.g. NOD-like receptor signalling and Toll-like receptor (TLR) signalling, were also upregulated upon HC/HF diet such as NOD-like receptor family, pyrin domain containing 3 (*NLRP3*) (p=0.007), *TLR2* (p<0.001) (**Figure 5C)**, and other genes (*TLR4*, *NOD2, PYCARD, CASP1*) **(Figure S5B-C**). Further, genes related to immune and T-cell signalling also showed increased gene expression upon HC/HF compared to HF/HC diet such as CD3 epsilon subunit of T-cell receptor complex (*CD3E*) (p=0.006) and integrin subunit alpha X (*ITGAX*) (p<0.001) (**Figure 5D**). Genes coding regulators of NF-κB activity such as protein kinase C beta (*PRKCB*) (p<0.001) and mitogen-activated protein kinase 1 (MAPK1) (p=0.001), also increased the daily expression levels upon HC/HF diet (**Figure 5E**). Finally, genes involved in reactive oxygen species (ROS) production and macrophage activation (*RAC2*, *PAK1*) were also upregulated in the HC/HF diet (**Figure S5D**).

**Figure 5.**
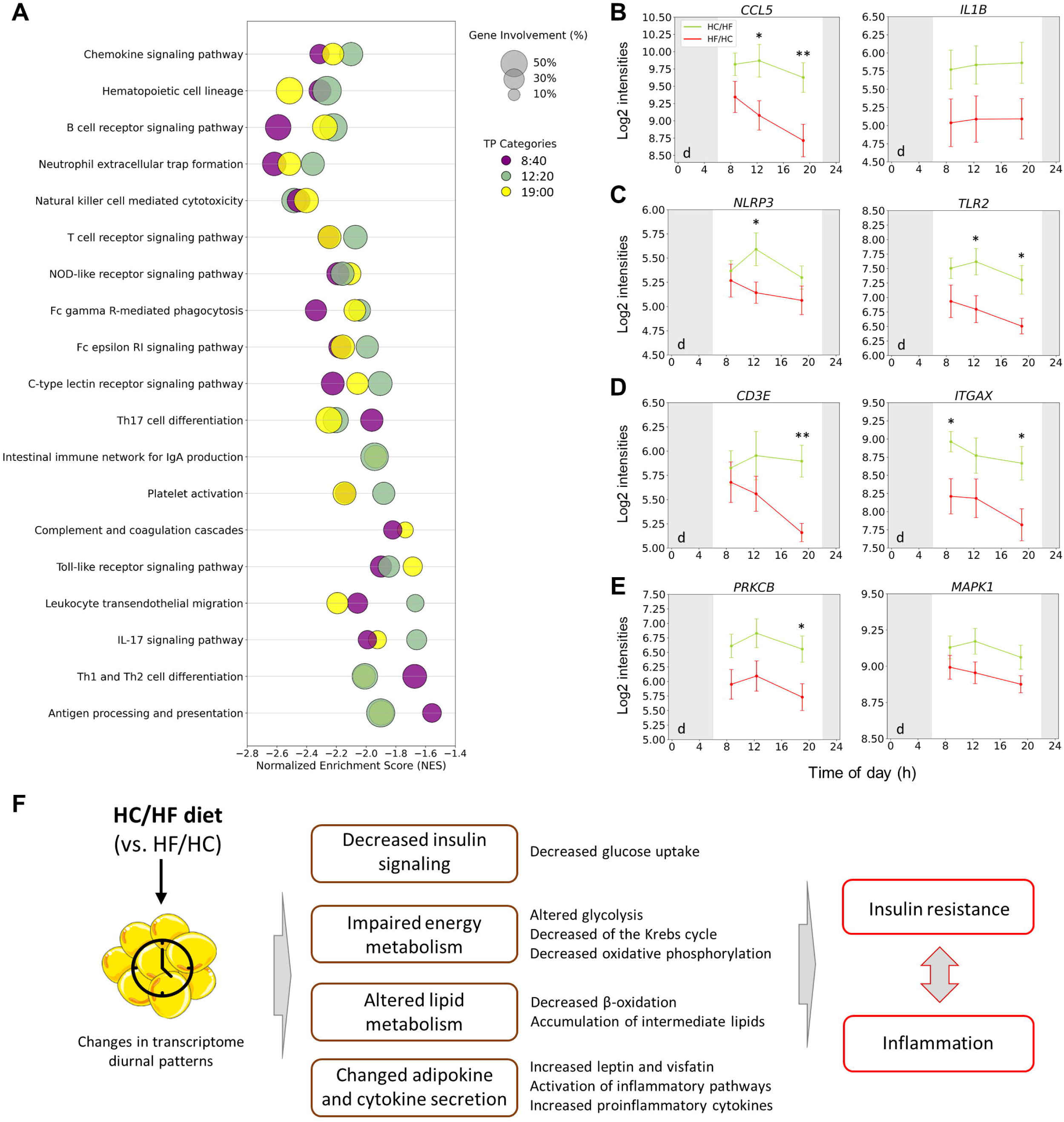
Timing of the carb and fat intake markedly changes inflammatory signaling in SAT. (A) Bubble plot representing inflammatory response pathways changed at 8:40 (purple), 12:20 (green), and 19:00 (yellow) after the HC/HF and HF/HC diets as revealed by GSEA. (B) Diurnal expression patterns of genes involved in inflammatory and chemotactic signalling after the HC/HF (green) and HF/HC (red) diets. (C) Diurnal expression patterns of genes involved in NOD-like receptor signalling and Toll-like receptor signalling after the HC/HF (green) and HF/HC (red) diets. (D) Expression patterns throughout the day of genes involved in immune and T-cell signalling after the HC/HF (green) and HF/HC (red) diets. (E) Diurnal expression patterns of genes involved in NF-κB activity after the HC/HF (green) and HF/HC (red) diets. (F) The proposed pathways that are altered under the HC/HF diet and could be promoting an early proinflammatory context in SAT. Higher fat intake in the afternoon could lead a decreased insulin sensitivity, altering energy metabolism performance by increasing certain steps of glycolysis, decreasing acetyl-CoA entry into the citrate cycle and oxidative phosphorylation. In addition, the decrease in global lipid metabolism in SAT could be leading to an accumulation of intermediate lipids that potentiates the state of insulin resistance. This could lead to increased metabolic stress, stimulating the expression of proinflammatory genes in the SAT and changes in adipokine and cytokine secretion. (A-E) n=15. Data are expressed as mean ± SEM for each time point. *p*-values were calculated by two-way repeated-measures ANOVA (d, by diet; t, by time). Genes which did not show diurnal oscillations on one or both diets were presented, the full oscillation analysis are in **Supplementary Dataset 2**. * *p*<0.05, ** *p*<0.01 indicate significant difference between HC/HF diet vs. HF/HC diet at the same time of the day by paired t-test. HC, high carbohydrate; HF, high fat; GSEA, gene set enrichment analysis.

### Timing of the carb and fat intake alters metabolic but not inflammatory gene expression in PBMC

The genes and signaling pathways altered by diets in SAT were additionally evaluated in PBMCs. Among clock genes, the *BMAL1* gene expression displayed no oscillation, showing a diet*time interaction effect (p=0.011) by two-way ANOVA (**Figure S6A**). The *PER1* gene exhibited rhythmic expression upon both diets. However, the *PER1* acrophase was affected by diurnal distribution of macronutrients, resulting in an advanced peak time upon HF/HC diet (p=0.008) (**Figure S6A**, **Table S4**). The *NR1D1* gene did not show changes depending on diet (**Figure S6A**). Similar to SAT, the metabolic genes *IRS1*, *IRS2* and *PDK4* showed diurnal oscillations in PBMCs upon one or both diets. No diet-dependent changes were observed in the diurnal oscillations of *IRS1*, while the *IRS2* gene only displayed oscillation upon HC/HF diet (**Figure S6B, Table S4**). Interestingly, the *PDK4* gene displayed a phase shift and a change in amplitude by diet (p=0.001 and p=0.014, respectively) (**Figure S6B, Table S4**), whereas the *PDK4* oscillatory patterns and dietary-induced changes were similar to SAT (**Figure 5B**). For inflammatory genes *CCL5*, *PRKCB, NLRP3, TLR2* and *IL1B* no expression differences between the diets were found (**Figure S6C**).

To test whether dietary-induced inflammatory changes in SAT can be observed in circulation, we further assessed circulating levels of interleukin-6 (IL6), the cytokine monocyte chemoattractant protein 1 (CCL2) and C-reactive protein (CRP) (**Figure S6D**). Consistent with the published data^37–39^, IL-6 showed an upward trend throughout the day and was oscillating in both diets (**Figure S6D**, **Table S4**). Serum levels of the inflammatory markers CCL2 and CRP showed no dietary effect and no circadian oscillations (**Figure S6D**).

## Discussion

Many studies in both humans and animals have demonstrated that dietary patterns, including meal composition and the intake timing of different nutrients, influence biological clocks as well as various physiological and metabolic processes^10,26,40–42^. However, while the ability of diet to modulate the oscillatory transcriptome profile of different tissues has been reported in animal models^26–28^, the extent of transcriptomic remodeling in humans remains unknown. This study investigated, for the first time in humans, the effects of a diurnal distribution of carb and fat intake on the oscillating and non-oscillating AT transcriptome. We compared the effects of two diurnal macronutrient patterns on the circadian clock, glucose and lipid metabolism and inflammatory response in SAT and additionally evaluated these pathways in PBMCs. We found that the change of the composition of morning and afternoon meals induces deep remodeling of the SAT transcriptome which is partly reflected in PBMCs.

White AT demonstrates strong circadian variations of metabolic function^33^. During the active phase, nutrient energy is stored in AT in the form of triglycerides, while during the inactive phase, which is usually accompanied by a fasting period, triglycerides are mobilized from AT stores and released as FFA, serving as energy substrates for other organs^43^. In this study, we confirmed the data reported by other authors in mice and in humans, that lipogenesis genes (e.g. *SREBF1*, *DGAT2* and *ACLY*) usually peak at the end of the active phase (evening in humans), and fatty acid oxidation genes (e.g. *PDK4*, *GADD45B* and *SESN1*) peak at the end of the resting phase (morning in humans)^7,33,44,45^. Similarly, key glucose metabolism genes in SAT, involved in the insulin signaling pathway (e.g. *IRS1, IRS2, PCK1*), phosphoinositide metabolism, glycolysis, gluconeogenesis, and pyruvate oxidation showed diurnal oscillations, consistent with recent studies^26,33^. In addition, a range of inflammatory genes involved in lymphocyte activation, differentiation, and cytokine production were also regulated in an oscillatory manner. In line with this, we previously demonstrated that circulating levels of the adipokines leptin and adiponectin, as well as the cytokine IL-6 exhibited diurnal variation in humans^25^.

Interestingly, the circadian transcriptome in SAT exhibited high plasticity and underwent remodelling in response to the nutrient challenge in our study^46^. This reprograming of diurnal oscillations in AT transcriptome has been previously reported in mice fed with a HF diet compared to those fed with a normal diet, highlighting the plasticity of AT in response to different dietary patterns^26^. In our study, we identified four groups of oscillating genes in human SAT: 1) genes showing the same oscillations after both diets; 2) genes changing oscillations after HF/HC vs HC/HF diet; 3) genes oscillating in the HC/HF diet but losing circadian rhythmicity in the HF/HC diet; 4) genes oscillating in the HF/HC diet but losing circadian rhythmicity in the HC/HF diet. Additionally, we observed an overall shift in peak times of up to four hours in oscillating genes, depending on the diurnal distribution of fat and carbs. Notably, studies in animal models have reported oscillatory gene expression remodelling in different tissues when comparing diets that differ in caloric intake and total composition (e.g. HF diet vs. normal diet)^26,28^ or the intake timing of a particular meal/nutrient in a control diet^47,48^. In this study, we provide the first evidence in humans that two isocaloric diets, differing in their distribution of macronutrients throughout the day, can imprint distinct gene expression signatures in SAT, changing their rhythmicity.

Many of genes oscillating upon both diets, which play a pivotal role in energy metabolism, showed alterations in amplitude or phase shift. In this regard, the *PDK4* gene is involved in metabolic plasticity as it inhibits pyruvate dehydrogenase complex and glucose oxidation, and promotes FFA oxidation, working as an energy substrate switch^49^. Diurnal oscillation of the *PDK4* gene expression has also been described in human SAT, in which it is observed that the maximum expression occurs in the early morning, at the end of the resting phase^50^. In our study, although both dietary patterns showed an acrophase close to early in the morning, the diurnal distribution of macronutrients promoted a phase shift. This effect could be due to the fact that the higher available exogenous glucose in the HF/HC diet during the afternoon prolongs lipogenesis, delaying the onset of lipid oxidation, lipolysis and gluconeogenesis that occur during the resting phase. In contrast, the higher available exogenous lipid and the lower exogenous glucose in the HC/HF diet during the afternoon stimulates the onset of lipid oxidation, lipolysis and gluconeogenesis, and inhibits lipogenesis earlier. Additionally, the correlation between serum FFA levels and the *PDK4* gene has also been reported, pointing out that the serum FFA levels regulate the expression pattern of this gene^50^. In fact, our results also showed that serum FFA levels were higher in the evening in response to the HC/HF diet and in the morning in response to the HF/HC diet^18^. Interestingly, the expression pattern of this gene was also affected by diet in PBMCs, showing a similar shift in the acrophases. This suggests that this metabolic effect would occur not only in SAT, but also in various peripheral tissues.

Other oscillating genes related to energy metabolism in SAT were also affected in this line. In fact, the *IRS1*, which is implicated in insulin-induced glucose uptake in AT^51^, and the *PCK1*, which is involved the glycerol-3-phosphate from non-glycidal precursors for fatty acid re-esterification in AT^52^, showed higher amplitudes after the HF/HC diet compared to the HC/HF diet. The diurnal patterns of *IRS1* and *IRS2* were also affected by diurnal distribution of macronutrients in PBMCs, although slightly different results might suggest tissue-specific changes^53^. Our results suggest that the different serum levels of postprandial glucose, lipids and hormones promote changes in the expression patterns of several energy metabolism pathways, leading to a more pronounced oscillation under the HF/HC diet. We hypothesized that the consumption of the HF food in the afternoon/evening (in combination with HC meals in the morning) reprograms the metabolic and inflammatory pathways in AT decreasing its insulin sensitivity. Indeed, HOMA-IR showed no changes of the whole-body insulin sensitivity, whereas AT-specific indices rQUICKI and the adiponectin/leptin ratio throughout the day revealed higher insulin sensitivity under HF/HC diet^29,30^. In line with this hypothesis, we observed higher gene expression over the day of non-oscillatory genes implicated in insulin-dependent glucose uptake pathways like *INSR* and *SLC2A4/GLUT4* in HF/HC diet, while *SLC2A1/GLUT1*, which is related to glucose transport in the basal state, was upregulated under HC/HF diet. Increased *GLUT1* transporter is associated with metabolic stress in other peripheral tissues^54^. Thus, the higher expression of *SLC2A1/GLUT1* could suggest a compensatory mechanism due to a possible lower insulin sensitivity upon the HC/HF diet. Additionally, energy metabolism was also affected, as only *HK1* and *PKM* showed higher expression upon the HC/HF diet, suggesting an increase in certain steps of glycolysis. However, the increased expression of *PDHA1* together with *UQCRC2* and *NDUFB5* under HF/HC diet suggests a lower transformation of pyruvate to acetyl-CoA, lower mitochondrial activity and lower ATP production through oxidative phosphorylation under HC/HF diet. Interestingly, it has been suggested that increased *HK1*, accompanied by a dissociation of mitochondrial activity, may contribute to type 2 diabetes-related disorders^55^. Indeed, GSEA revealed decreased citrate cycle, pyruvate metabolism, glycolysis/gluconeogenesis and oxidative phosphorylation upon the HC/HF diet compared to the HF/HC diet. These findings would reinforce the hypothesis that higher postprandial lipid levels in the afternoon would result in lower insulin sensitivity, decreasing the amplitude of the diurnal oscillation and the expression pattern along the day of the different insulin-dependent metabolic pathways.

Further, insulin, one of the most important meal-induced hormones, can entrain peripheral clocks and clock-controlled genes and affect the huge network of metabolic pathways in insulin-sensitive tissues, such as liver and AT^56–58^. Because meal-induced hormone secretion is strongly dependent on both the composition of the meal and the time of intake^17,18^, different metabolic responses should be expected if the same meal is ingested in the morning or in the evening. This could also explain the higher expression throughout the whole day in non-oscillatory genes under HF/HC diet in genes involved in energy metabolism in SAT. It could also be hypothesized that a dietary pattern consumed over a long period of time may predetermine the level of gene expression during the whole day. In addition, the gain of oscillatory gene expression depending on diurnal dietary patterns could be due to a compensatory mechanism in AT in response to time-of-day variations in macronutrients^59^. However, further research is needed to elucidate exact metabolic mediators and points of regulation, as well as time of a dietary pattern required for the onset of the remodelling of AT transcriptome.

Beside the remodelling of metabolic pathways, the second most important finding of our study was the large dietary effect on the tissue inflammation. Indeed, we unexpectedly found dietary effects on various aspects of inflammatory response including T cell, B cell and macrophage activation, adhesion and migration, cytotoxicity, phagocytosis and cytokine production. Such proinflammatory changes characterized by the secretion of proinflammatory cytokines and infiltration of proinflammatory immune cells are described for obesogenic concept due to the excessive fat storage^60,61^. Activation of key inflammatory pathways such as NF-κB, JNK and NLRP3 inflammasome, as well as the excess of FFAs which can be converted into lipid intermediates such as diacylglycerols and ceramides downregulate insulin signaling and favor a type 2 diabetes scenario^35,62^. Similar changes were also shown upon the diet rich in saturated fats even when total caloric intake is held constant although carefully controlled human trials and findings in human AT are limited^63–65^. Surprisingly, in our study, many genes involved in the abovementioned inflammatory-related mechanisms showed a higher expression under the HC/HF diet compared to HF/HC diet, meaning that even specific timing of the high-fat consumption can favor AT inflammation. This finding is in consistence with our previous data about the effect of the same diets on the LPS-induced cytokine secretion in blood lymphocytes^25^. We hypothesize that the HC/HF diet characterized by higher fat intake in the afternoon, could remodel the diurnal signature in AT promoting the inflammation via many factors as discussed below.

First, we observed dietary-induced alterations of the serum leptin and visfatin (eNAMPT) levels^25^, adipokines involved the regulation of inflammatory response and typically associated with obesity^66^. In fact, the gene expression of the *NAMPT* gene in SAT was higher in HC/HF diet compared to HF/HC diet, especially at 19:00. Although this gene exhibits anti-inflammatory functions in AT, NAMPT also acts as an adipokine when it is secreted extracellularly (visfatin/eNAMPT) and contributes to chronic inflammatory diseases^67,68^. This is in line with the higher serum visfatin levels at 19:00 under the HC/HF diet compared to the HF/HC diet that we reported previously^25^. Additionally, serum leptin also showed higher levels during the day in response to the HC/HF diet whereas adiponectin remained similar in both diets ^25^. This is in consistence with data from mice that consumption of HF food at the end of active period induces hyperleptinemia^20^, and with the increased expression of key inflammatory genes during the day found in our data. Secondly, leptin is able to activate the production of pro-inflammatory cytokines IL-6, TNF-α, and IL-12 in monocytes and macrophages and of pro-inflammatory Th1 cytokines in CD4^+^ T cells^69,70^, and these effects might be time-dependent. Although, we did not find dietary effects on circulating levels of inflammatory markers IL-6 and CRP^25^ or on the expression of inflammatory genes in PBMCs in the same subjects, tissue-specific effects of the diets on the inflammatory state in SAT might occur. Thirdly, ceramides were one of the lipid metabolites increased after MTT-HF that we reported in previous studies^71^. This metabolite is not only associated with insulin resistance but also with inflammation as it has been shown to promote p38 MAPK and JNK signalling pathways^72^. In addition, this metabolite also stimulates IL-1B secretion and activates NLRP3 in macrophages^73^. Accordingly, our results showed increased expression of *NLRP3* and *IL1B* and other genes implicated in the development of AT inflammasome like *TLRs*.

Taking together, our findings suggest that the HC/HF dietary pattern could be promoting an early proinflammatory context and associated insulin resistance in SAT, possibly due to a higher fat intake in the afternoon. Timely remodelling of glucose and lipid metabolism induced by HC/HF diet^71^ could decrease the AT-specific insulin sensitivity^35,62^, which was accompanied by alterations of energy metabolism with increasing certain steps of glycolysis, decreasing acetyl-CoA entry into the citrate cycle and oxidative phosphorylation^55^. This could lead to higher metabolic stress, increasing *NAMPT* expression and circulating levels of visfatin and leptin and stimulating the expression of proinflammatory genes in SAT (**Figure 5F**). Whether inflammatory pathways are activated by metabolic changes or, vice versa, insulin sensitivity induced by inflammation and how exactly the timing of the fat and card intake induce both alterations need to be clarified in preclinical studies.

Remarkably, core clock genes in SAT and salivary cortisol used as a central clock marker showed diurnal variations under both diets, and overall, their parameters were not generally affected by different patterns of meal composition. This confirms that the central clock and core clock genes in SAT, are highly resistant to nutrient-induced alterations, whereas clock output genes are more sensitive to nutrient challenge as demonstrated by Damiola *et al*.^74^. Importantly, in our study, only meal composition during the daytime was modulated without any shift of the usual food intake window, therefore no large effects of diet on serum cortisol and on clock genes in the SAT are to be expected. In fact, diurnal variation in insulin sensitivity (modulated by lipid and glucose levels throughout the day) may be sufficient to promote changes in circadian-regulated metabolic mechanisms but not to SAT core clock genes under these isocaloric-diet conditions. However, significant changes were observed in the oscillatory pattern of *BMAL1*, *PER1* and *NR1D1* in PBMCs. In fact, although neither diet showed any significant oscillation in *BMAL1* expression, the expression pattern was opposite between the two diets. This was accompanied by a change in *PER1* acrophase. The different observations in SAT and PBMC might reflect tissue-specific differences. Indeed, we previously reported altered oscillations of core clock genes in CD14+ monocytes upon whole-day isocaloric HF diet compared to the HC diet^31^. Furthermore, these results suggest a possible alteration in the synchronisation of the global clock system as the composition of the diet may be influencing the clock of some peripheral tissues more markedly than in SAT^36^.

Some limitations of our study should be mentioned. Firstly, the analysis of diurnal patterns in our study is based on three SAT samples collected throughout the day time, but not in the night time, because 24-hour sampling was not feasible for technical reasons. To address this limitation, we used a modified approach applying a magnitude correction and the iterative algorithm, as described in detail previously^31^. This approach was previously validated vs. 24-hour sampling and allows analysis of oscillation parameters with reasonable accuracy, given a good ratio from amplitudes to noise. Nevertheless, this method is unable to detect or accurately characterize oscillations with a low amplitude. This can explain why the number of oscillating genes detected in our study (4.7-4.9%) is lower than in similar published human studies in SAT using 24-hour sampling (8.4%)^75^. However, we were able to detect diurnal oscillations both in core clock and in key metabolic and inflammatory genes described previously^33,75,76^. This confirms that our approach can deliver reliable results and be beneficial for studying diurnal oscillations in the clinical setting when 24-hour sampling is not feasible. Secondly, the study was conducted in overweight, non-diabetic men, so it is not possible to extrapolate these effects to women or other population groups given the sex-dependent^77,78^ or metabolic status-dependent^18,79^ differences found in the clock system reported by other authors. Therefore, further studies in other population groups are needed to confirm these findings.

In conclusion, our study investigated for the first time in humans the effects of isocaloric temporal segmentation of carbohydrate and fat intake on the AT transcriptome. High-throughput profiling revealed that different dietary patterns induce a deep remodeling of the AT transcriptome in humans, changing the diurnal oscillatory signature of transcriptome expression. These changes affected glucose and lipid metabolism and inflammatory pathways, suggesting that a diet with higher fat intake in the afternoon may promote an early proinflammatory state in SAT. Our findings contribute to a better understanding of chrononutrition for the development of new strategies to prevent the metabolic disturbances and systemic inflammation that characterize obesity and type 2 diabetes.

## Data Availability

All data produced in the present study are available upon reasonable request to the authors.

## Acknowledgements

We thank all study participants for their cooperation. We gratefully acknowledge the excellent technical assistance of Katrin Sprengel, Tanja Ahrens, Andrea Borchert, Alexandra Ullrich, Melanie Hannemann, Stephanie Peglow, and Dominique Zschau. We thank Stephanie Sucher for her extensive advice regarding nutritional counselling. We also thank all students for their help in sample or data analysis. We acknowledge Dr. Celine Vetter und Prof. Till Roenneberg (Centre for Chronobiology of Institute of Medical Psychology of University of Munich, Germany) for the analysis of MCTQ data.

## Author contributions

KK, AFHP, and OPR designed the research; KK and SH conducted the research; JRS-R, KJ, CS, and OPR analyzed data; JRS-R, KK, AK, AFHP, and OPR wrote and edited the manuscript; OPR has primary responsibility for the final content. All authors read and approved the final manuscript.

## STAR Methods

### KEY RESOURCES TABLE

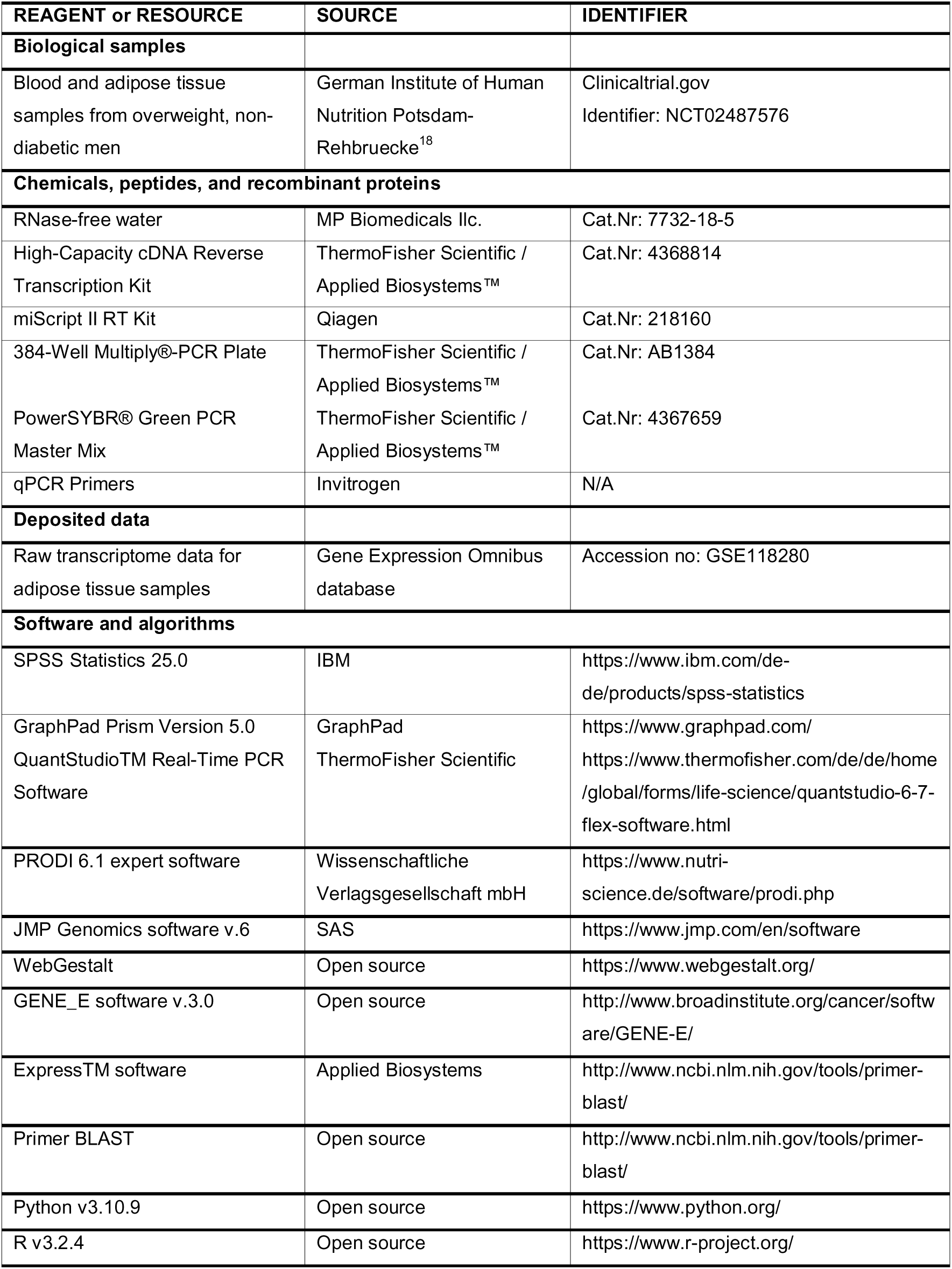

### RESOURCE AVAILABILITY

#### Lead contact

Further information and requests for resources and reagents should be directed to and will be fulfilled by the lead contact, Olga Pivovarova-Ramich.

#### Materials availability

This study did not generate new unique reagents.

#### Data and code availability

- Transcriptome data supporting the findings of this study are included within the Article and Supplemental Materials. The access number for this dataset is listed in the key resources table.
- This paper does not report original code.
- Any additional information required to reanalyse the data reported in this paper is available from the lead contact upon request.

### EXPERIMENTAL MODEL AND STUDY PARTICIPANT DETAILS

#### Subjects and design of the study

Details of study design, recruitment and examination of study participants and dietary interventions were previously published^18,25,71,80^. Briefly, 32 overweight men without diabetes were enrolled, and 29 men completed the cross-over study (**Figure S1**). We compared effects of two 4-week isocaloric diets – (1) HC/HF diet consisted of a high-carb breakfast and lunch (65 EN% carbohydrates, 20 EN% fat, 15 EN% protein) consumed between 6:00 h and 13:30 h and a high-fat snack and dinner (35 EN% carbohydrates, 50 EN% fat, 15 EN% protein) consumed between 16:30 h and 22:00 h and (2) HF/HC diet consisted of a high-fat breakfast and lunch and a high-carb snack and dinner, with a 4-week washout phase between intervention periods. At the end of each intervention period, two standardized meal tolerance tests, high-carb (835 kcal, 64.8 EN% CHO, 14.8 EN% protein, 20.3 EN% fat) or high-fat (849 kcal, 35.3 EN% CHO, 15.1 EN% protein, 49.6 EN% fat), were performed at 09.00 h and 15.40 h during the investigation day according to each participant’s previous diet^18^. Gastric emptying rate was determined by the ^13^C-acetate breath test as described^18^.

SAT samples were collected three times during the investigation day (at 8.40, 12.20 and 19.00 h) at the level of the umbilicus by the needle aspiration as described^71^. Peripheral blood mononuclear cells (PBMC) were isolated by gradient centrifugation with Ficoll at the same day time (at 8.55, 12.00 and 19.00 h) as described^31^. Saliva samples were collected using Salivette Collection Device (Salivettes, Sarstedt, Germany) one day before each clinical investigation day (CID) every 4 h during 24h. Individual chronotypes and sleep/wake times were assessed using the Munich Chronotype Questionnaire (MCTQ)^81^. Dietary compliance was assessed by analysis of food records completed for five consecutive days prior to both investigation days with PRODI 6.1 expert software (Nutriscience, Germany)^25^. The study was conducted according to the Declaration of Helsinki, approved by the Ethics Commission of Charité University Medicine, Berlin, Germany (EA2/074/12) and registered at www.clinicaltrials.gov (NCT02487576). All study participants provided oral and written informed consent.

### METHOD DETAILS

#### Measurement of biochemical markers

Serum glucose levels and free fatty acids (FFA) and C-reactive protein (CRP) were measured using ABX Pentra 400; (HORIBA, ABX SAS, France). Commercial enzyme-linked immunosorbent assays (ELISA) were used for measurement of serum insulin (Mercodia, Sweden), IL-6 and monocyte chemoattractant protein-1 (CCL2/MCP-1) (both from BioTechne R&D Systems GmbH, Germany), and salivary melatonin and cortisol (IBL, Germany) as described previously^18,25^.

#### RNA isolation

SAT samples of 15 randomly selected participants were processed for the transcriptome analysis. PBMC samples of 29 participants were processed for qPCR analyses. Total RNA was purified using the miRNeasy Lipid Tissue Mini Kit (Qiagen, Germany) for SAT samples and by NucleoSpin RNA Kit (Macherei-Nagel, Germany) for PBMC samples. RNA quality was assessed by Agilent 2100 bioanalyzer using Agilent RNA 6000 Nano Kit (Agilent Technologies, Germany). RNA concentration was measured using an ND-1000 spectrophotometer (Nanodrop, PeqLab).

#### Transcriptome analysis

Single-stranded cDNA was synthesized from the SAT RNA with miScript II RT Kit (Qiagen, Germany). Biotinylated cRNA were prepared according to the standard Affymetrix protocol from 300 ng total RNA and hybridized to GeneChip Human Gene 2.0 ST Arrays (Affymetrix, Germany) for 16h at 45° C. Microarrays were scanned with an Affymetrix GeneArray Scanner 3000. CustomCDF Version 14 with Entrez based gene definitions was used for gene annotation. The Raw fluorescence intensity values were normalized applying quantile normalization, RMA background correction and Medianpolish Probeset Summary. Microarray data were analysed with JMP Genomics software v.6 (SAS, USA). Functional annotation was done with WebGestalt^82^. Gene set enrichment analysis (GSEA) was performed using gseapy package in Python v3.10.9. Heatmaps were created using GENE_E software v.3.0. Microarray data were deposited in Gene Expression Omnibus database (accession no. GSE118280).

#### Quantitative real-time PCR (qPCR)

Single-stranded cDNA was synthesized from PBMC RNA with High-Capacity cDNA Reverse Transkription Kit^™^ (Applied Biosystems, Germany). The qPCR was performed by ViiA 7 sequence detection system using Power SYBR Green PCR Master Mix (Applied Biosystems, USA). Quantification of mRNA levels was made by the standard curve method. Gene expression was normalized to the reference gene *GUSB* (beta-glucuronidase) and *B2M* (Beta-2-microglobulin). Primers were designed with Primer ExpressTM software (Applied Biosystems, USA) or Primer BLAST and are listed in **Table S5**.

### QUANTIFICATION AND STATISTICAL ANALYSIS

#### Analysis of circadian rhythms of gene expression and salivary markers

Parameters of diurnal oscillations of gene expression and salivary markers were estimated by a three-time-point rhythm prediction method established previously^31^. In brief, genes demonstrating a time or time*diet effects in two-way repeated measures ANOVA (p<0.01) were analysed to predict the 24h rhythm. For this, sinus models were fitted to the gene expression data of all donors to obtain values for amplitude, mesor and peak time:

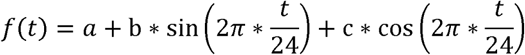

The mesor was defined as mean of the oscillation, amplitude – as half of the distance between the peak and trough, and peak time (acrophase) - as time of the maximum of the fitted curve. In present paper, we used a modified approach applying a magnitude correction and the iterative algorithm. This correction was performed by calculating the mean of the gene expression values per donor and per diet, in order to minimize inter-donor variability in absolute expression levels and better estimation of oscillatory patterns. The magnitude correction allowed to achieve higher r2-values and lower p-values upon the sinus fitting of the three-time-point data compared with previously published method^31^ and to detect more genes showing diurnal rhythms. Because of magnitude correction, our novel approach allowed to compare amplitude and acrophases of revealed rhythmic genes but not their mesors. Significant differences in oscillations between the two diets were carried out with the *compareRhythms* package^32^, a package specifically for oscillatory analysis of transcriptomics data (amplitude > 0.2, p <0.05 and FDR <0.1). Differences between circadian parameters (amplitude and acrophase) were analyzed using the CosinorPy package (p<0.05)^83^.

Because of the more complex nature of the salivary cortisol rhythm including awakening response, an extended sinus model was used for fitting cortisol data:

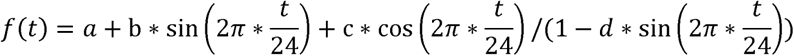

For cortisol, a nadir (time of the curve minimum) was calculated additionally as a more robust marker of phase of central clock in humans compared with cortisol acrophase^84^. Significance of fitted models was compared to corresponding constant models using an F-test. For salivary melatonin, we were not able to find a fitting model^34^ providing a reasonable accuracy of diurnal rhythm analysis.

#### Statistical analysis

Statistical analyses were performed with JMP Genomics software v.6 (SAS, USA) and SPSS v.20 (IBM, USA). SAT genes, serum parameter and salivary parameters data were presented as means ± SEM per time point. Time and diet effects were estimated using two-way repeated measures ANOVA, with p<0.01 considered statistically significant. Post-hoc comparisons between the two diets at each time point were performed using paired Student’s t-test. Multiple correction for p values was calculated using the Benjamini–Hochberg method of False Discovery Rate (FDR) control for two-way repeated measures ANOVA and circadian analysis comparison with the *compareRhythms* package. Parameters of circadian oscillations (the amplitude and the peak time) were estimated by a three-time-point rhythm prediction method^31^ with a magnitude correction using R software v.3.2.4. Data were plotted using GraphPad Prism Version 5.0 and Python v3.10.9.

### ADDITIONAL RESOURCES

The study was registered at www.clinicaltrials.gov (NCT02487576).

