## Supplementary Material for "Remodeling of human diurnal adipose tissue transcriptome by the composition of morning and afternoon meals"

**Content	Page**

**Supplemental tables**

**Table S1. Clinical characteristics of study subjects.	3**

**Table S2. Functional annotation of oscillating genes by KEGG pathway analysis.	4**

**Table S3. Oscillating genes showed a dietary effect on the peak time or amplitude.	7**

**Table S4. Oscillation parameters of transcripts in SAT and PBMC and biomarkers in saliva and serum.	11**

Table S5. Primers used for real-time PCR.	12

**Supplemental figures**

**Figure S1.** Study flow chart.	13

**Figure S2.** Adiponectin/leptin ratio and gastric emptying rates after meal tolerance tests (n=15).	14

**Figure S3.** Meal composition change the transcriptome profile in adipose tissue at 19:00 time point (n=15).	15

**Figure S4. Oscillating and non-oscillating genes involved in lipid and glucose metabolism in SAT (n=15).**	16

**Figure S5. Oscillating and non-oscillating genes involved in inflammatory response in SAT (n=15).	18**

**Figure S6.** Diet composition influenced diurnal oscillations and expression levels of glucose and lipid metabolism genes in PBMCs (n=15).	19

**Table S1. Clinical characteristics of study subjects.**

|  | **HC/HF diet** | **HF/HC diet** | **p-value** |
| --- | --- | --- | --- |
| N (%male) | 15 (100) | |  |
| NGT/IFG/IGT [n] | 8 / 5 / 3 | |  |
| Age [years] | 44.9 ± 3.7 | |  |
| Weight [kg] | 88.3 ± 4.6 | 88.6 ± 4.6 | 0.313 |
| BMI [kg/m²] | 27.4 ± 1.1 | 27.6 ± 1.1 | 0.180 |
| Waist circumference [cm] | 92.4 ± 3.1 | 94.3 ± 3.0 | 0.031 |
| Total body fat [%] | 24.5 ± 2.1 | 24.8 ± 2.2 | 0.561 |
| Total cholesterol [mmol/l] | 4.84 ± 0.23 | 4.84 ± 0.23 | 0.962 |
| HDL cholesterol [mmol/l] | 1.09 ± 0.05 | 1.07 ± 0.05 | 0.386 |
| LDL cholesterol [mmol/l] | 3.20 ± 0.20 | 3.21 ± 0.21 | 0.897 |
| Triglycerides [mmol/l] | 1.22 ± 0.19 | 1.22 ± 0.19 | 0.892 |
| NEFA [mmol/l] | 0.48 ± 0.02 | 0.46 ± 0.03 | 0.660 |
| HbA1c [%] | 5.27 ± 0.07 | 5.37 ± 0.10 | 0.360 |
| Fasting glucose [mmol/l] | 5.47 ± 0.11 | 5.43 ± 0.10 | 0.601 |
| Fasting insulin [pmol/l] | 42.6 ± 4.6 | 41.1 ± 7.0 | 0.718 |
| HOMA-IR [mmol*mU *l^-2^] | 1.51 ± 0.18 | 1.46 ± 0.27 | 0.745 |
| rQUICKI | 0.42 ± 0.01 | 0.45 ± 0.02 | **0.044** |
| Daily calorie intake [kcal]^a^ | 2859 ± 148 | 2759 ± 112 | 0.375 |
| Dietary intake,  6:00-13:30 (carbs/fat/protein, EN%) | 65.6 / 20.2 / 14.2 | 35.3 / 49.4 / 15.3 |  |
| Dietary intake,  16:30-22:00 (carbs/fat/protein, EN%) | 34.2 / 50.6 / 15.2 | 64.8 / 20.8 / 14.3 |  |
| Chronotype [MSF-Sc] | 3.33 ± 0.26 | 3.40 ± 0.24 | 0.508 |
| Sleep offset [hh:mm]^b^ | 05:59 ± 0:15 | 06:10 ± 0.17 | 0.244 |
| Sleep onset time [hh:mm]^b^ | 22:53 ± 0:16 | 22:56 ± 0:21 | 0.664 |
| Sleep duration [min]^b^ | 425 ± 10 | 435 ± 13 | 0.423 |

Data were collected at the end of each intervention (visit 2 and 4). ^a^ – data on food intake collected during 14 days of intervention; ^b^ – sleep times and duration on working days at the end of each intervention. Data are shown as mean ± SEM. MSF-Sc, mid-sleep time point on free days adjusted for individual average sleep need accumulated on work days determined using MCTQ; NGT, normal glucose tolerance; IFG, impaired fasting glucose; IGT, impaired glucose tolerance; rQUICKI, revised quantitative insulin sensitivity check index.

**Table S2. Functional annotation of oscillating genes by KEGG pathway analysis.**

| Term | Gene No. | p-value | FDR | Name of genes |
| --- | --- | --- | --- | --- |
| Genes showing the same oscillations after both diets (995) | | | | |
| Circadian rhythm | 10 | 8.89E-07 | 2.81E-04 | BMAL1/CRY1/CRY2/DBP/NFIL3/NR1D2/PER1/PER2/PER3/RORB |
| Notch signaling pathway | 11 | 5.04E-05 | 0.008 | ADAM17/DLL1/HES1/HEY2/JAG1/LOC102723796/MFNG/NOTCH1/NOTCH4/NRARP/SPEN |
| AGE-RAGE signaling pathway in diabetic complications | 12 | 0.001 | 0.119 | AGTR1/BCL2/CASP3/CCL2/FOXO1/ICAM1/IL6/PLCD1/PRKCD/SELE/THBD/VEGFC |
| p53 signaling pathway | 9 | 0.004 | 0.239 | BCL2/CASP3/CCND3/GADD45B/RCHY1/SESN1/SESN2/SESN3/THBS1 |
| Transcriptional misregulation in cancer | 17 | 0.004 | 0.239 | BCL6/CEBPB/DUSP6/FOXO1/GADD45B/H3F3B/HHEX/HIST1H3I/HOXA10/IL6/KLF3/MAF/PAX3/PER2/PLAU/SIX1/ZBTB16 |
| TNF signaling pathway | 12 | 0.005 | 0.239 | ADAM17/CASP3/CCL2/CEBPB/CHUK/CREB5/ICAM1/IL6/JAG1/MMP14/SELE/VEGFC |
| Parathyroid hormone synthesis, secretion and action | 11 | 0.010 | 0.397 | AKAP13/BCL2/CREB5/GNA11/HBEGF/KL/MMP14/MMP15/NACA/PLD1/SGK1 |
| Glycosaminoglycan biosynthesis - chondroitin sulfate / dermatan sulfate | 4 | 0.011 | 0.397 | CHST3/CHSY1/DSEL/UST |
| ABC transporters | 6 | 0.011 | 0.397 | ABCA10/ABCA6/ABCB1/ABCC5/ABCD2/ABCG1 |
| Autophagy - animal | 14 | 0.013 | 0.401 | ATG14/BCL2/DAPK1/DAPK2/DDIT4/DEPTOR/GABARAPL2/MTMR14/NRBF2/PRKCD/RAB7B/TBK1/TRAF6/VPS18 |
| Protein processing in endoplasmic reticulum | 14 | 0.014 | 0.401 | BAG2/BCL2/CKAP4/DERL2/DNAJB1/HSPA1B/HSPBP1/PLAA/SAR1B/SEC61A2/SYVN1/UBQLN4/UBXN6/XBP1 |
| Insulin resistance | 10 | 0.017 | 0.453 | CREB5/FOXO1/GYS1/IL6/PPARGC1A/PPARGC1B/PPP1R3B/PPP1R3C/PRKCD/SREBF1 |
| Alcoholic liver disease | 12 | 0.019 | 0.470 | ADH1B/ADH1C/C3AR1/CASP3/CHUK/FOXO1/IL6/LPIN3/PPARGC1A/SREBF1/TBK1/TRAF6 |
| Glycosphingolipid biosynthesis - ganglio series | 3 | 0.024 | 0.534 | ST3GAL1/ST3GAL5/ST6GALNAC3 |
| NF-kappa B signaling pathway | 9 | 0.034 | 0.690 | BCL2/BLNK/CHUK/EDA2R/GADD45B/ICAM1/IL1R1/PLAU/TRAF6 |
| Efferocytosis | 12 | 0.035 | 0.690 | ADAM17/CASP3/CEBPB/DUSP4/MERTK/PLA2G6/PTGER4/RAB7B/SGK1/SLC16A1/THBS1/VPS18 |
| TGF-beta signaling pathway | 9 | 0.040 | 0.740 | BAMBI/ID1/ID3/LEFTY2/RBL1/SKIL/SMAD6/SMAD7/THBS1 |
| IL-17 signaling pathway | 8 | 0.048 | 0.849 | CASP3/CCL2/CEBPB/CHUK/IL6/MAPK6/TBK1/TRAF6 |
| Genes changing oscillations after HF/HC vs HC/HF (277-199) | | | | |
| MAPK signaling pathway | 12 | 0.003 | 0.311 | AREG/CACNA2D3/DDIT3/DUSP10/DUSP7/KIAA1804/PDGFA/SOS2/SRF/STMN1/TGFBR1/VEGFA |
| Adipocytokine signaling pathway | 5 | 0.005 | 0.311 | CPT1A/IRS1/IRS2/PCK1/PPARA |
| FoxO signaling pathway | 7 | 0.006 | 0.311 | C10orf10/CCNG2/IRS1/IRS2/PCK1/SOS2/TGFBR1 |
| Hepatocellular carcinoma | 8 | 0.006 | 0.311 | AXIN2/BAK1/CDK6/FZD8/KEAP1/SOS2/TGFBR1/TXNRD1 |
| Type II diabetes mellitus | 4 | 0.007 | 0.311 | HK2/IRS1/IRS2/SOCS4 |
| Insulin resistance | 6 | 0.008 | 0.311 | CPT1A/IRS1/IRS2/MGEA5/PCK1/PPARA |
| RNA degradation | 5 | 0.008 | 0.311 | BTG2/CNOT8/EXOSC4/LSM1/LSM4 |
| Glycosylphosphatidylinositol (GPI)-anchor biosynthesis | 3 | 0.012 | 0.322 | PIGC/PIGM/PIGO |
| Cushing syndrome | 7 | 0.013 | 0.322 | AXIN2/CDK6/FZD8/KCNA4/LDLR/PLCB1/RASD1 |
| Colorectal cancer | 5 | 0.013 | 0.322 | AREG/AXIN2/BAK1/SOS2/TGFBR1 |
| Hepatitis C | 7 | 0.014 | 0.322 | BAK1/CDK6/LDLR/PPARA/PPP2CA/RIPK1/SOS2 |
| Endometrial cancer | 4 | 0.015 | 0.322 | AXIN2/BAK1/CTNNA3/SOS2 |
| Insulin signaling pathway | 6 | 0.024 | 0.455 | HK2/IRS1/IRS2/PCK1/SOCS4/SOS2 |
| Breast cancer | 6 | 0.032 | 0.455 | AXIN2/BAK1/CDK6/FZD8/HEYL/SOS2 |
| PPAR signaling pathway | 4 | 0.034 | 0.455 | CPT1A/HMGCS1/PCK1/PPARA |
| Glioma | 4 | 0.034 | 0.455 | BAK1/CDK6/PDGFA/SOS2 |
| Gastric cancer | 6 | 0.034 | 0.455 | AXIN2/BAK1/CTNNA3/FZD8/SOS2/TGFBR1 |
| Pancreatic cancer | 4 | 0.036 | 0.455 | BAK1/CDK6/TGFBR1/VEGFA |
| Chronic myeloid leukemia | 4 | 0.036 | 0.455 | BAK1/CDK6/SOS2/TGFBR1 |
| ATP-dependent chromatin remodeling | 5 | 0.041 | 0.455 | BRD8/CHD4/HIST1H2AJ/MORF4L2/MTA2 |
| Hippo signaling pathway | 6 | 0.041 | 0.455 | AREG/AXIN2/CTNNA3/FZD8/PPP2CA/TGFBR1 |
| Valine, leucine and isoleucine degradation | 3 | 0.042 | 0.455 | AACS/ALDH1B1/HMGCS1 |
| Polycomb repressive complex | 4 | 0.045 | 0.455 | BCOR/BCORL1/CBX4/YAF2 |
| AMPK signaling pathway | 5 | 0.047 | 0.455 | CPT1A/IRS1/IRS2/PCK1/PPP2CA |
| Growth hormone synthesis, secretion and action | 5 | 0.047 | 0.455 | GH2/IRS1/IRS2/PLCB1/SOS2 |
| Histidine metabolism | 2 | 0.048 | 0.455 | ALDH1B1/C9orf41 |
| Cholesterol metabolism | 3 | 0.049 | 0.455 | C19orf80/LDLR/MYLIP |
| Shigellosis | 8 | 0.049 | 0.455 | ACTR3B/ELMO2/HK2/PFN1/PLCB1/PLCD3/RIPK1/SHARPIN |

Genes demonstrated significant oscillations in the analysis by rhythm prediction method (p<0.05, amplitude >0.2) were subjected to pathway analysis identified by WebGestalt (<http://www.webgestalt.org/>) using the Kyoto Encyclopedia of Genes and Genomes (KEGG) database and the human genome as a background.

**Table S3. Oscillating genes showed a dietary effect on the peak time or amplitude.**

| **Gene** | **Diet** | **Cosinor fitting** | | **Amplitude** | **P (Amp)*** | **Acrophase (h)** | **P (Acr)*** |
| --- | --- | --- | --- | --- | --- | --- | --- |
|  |  | **r^2^** | **P*** |  |  |  |  |
| *AACS* | HC/HF | 0.343 | 1.48E-04 | 0.246 | 0.074 | 22:12 | 0.449 |
|  | HF/HC | 0.645 | 3.55E-10 | 0.398 |  | 20:43 |  |
| *ADAMTS4* | HC/HF | 0.749 | 2.44E-13 | 0.516 | **0.001** | 18:12 | 0.751 |
|  | HF/HC | 0.866 | 1.11E-16 | 0.822 |  | 17:57 |  |
| *AGAP5* | HC/HF | 0.415 | 1.30E-05 | 0.205 | 0.724 | 16:52 | **0.000** |
|  | HF/HC | 0.220 | 5.38E-03 | 0.238 |  | 01:06 |  |
| *AKIRIN1* | HC/HF | 0.825 | 1.11E-16 | 0.336 | **0.005** | 16:17 | **0.028** |
|  | HF/HC | 0.657 | 1.69E-10 | 0.203 |  | 17:59 |  |
| *ALDH1B1* | HC/HF | 0.258 | 1.91E-03 | 0.274 | 0.993 | 00:37 | **0.002** |
|  | HF/HC | 0.446 | 4.17E-06 | 0.273 |  | 19:24 |  |
| *AOC4P* | HC/HF | 0.178 | 1.61E-02 | 0.503 | 0.209 | 14:10 | 0.061 |
|  | HF/HC | 0.557 | 3.71E-08 | 0.762 |  | 13:06 |  |
| *ATOH1* | HC/HF | 0.228 | 4.31E-03 | 0.206 | 0.874 | 11:34 | **0.000** |
|  | HF/HC | 0.122 | 6.55E-02 | 0.227 |  | 02:22 |  |
| *AXIN2* | HC/HF | 0.287 | 8.26E-04 | 0.348 | 0.625 | 15:44 | 0.052 |
|  | HF/HC | 0.579 | 1.29E-08 | 0.412 |  | 17:56 |  |
| *BCOR* | HC/HF | 0.333 | 2.04E-04 | 0.263 | 0.788 | 15:46 | **0.019** |
|  | HF/HC | 0.481 | 1.03E-06 | 0.239 |  | 19:19 |  |
| *BHLHE40* | HC/HF | 0.262 | 1.69E-03 | 0.347 | 0.250 | 14:21 | **0.001** |
|  | HF/HC | 0.622 | 1.37E-09 | 0.485 |  | 16:12 |  |
| *C10orf10* | HC/HF | 0.404 | 1.90E-05 | 0.949 | 0.781 | 03:37 | **0.033** |
|  | HF/HC | 0.826 | 1.11E-16 | 0.887 |  | 05:05 |  |
| *C19orf80* | HC/HF | 0.405 | 1.84E-05 | 0.238 | **0.000** | 21:34 | 0.604 |
|  | HF/HC | 0.636 | 6.22E-10 | 0.628 |  | 20:16 |  |
| *C8orf4* | HC/HF | 0.411 | 1.50E-05 | 0.299 | 0.064 | 22:26 | 0.169 |
|  | HF/HC | 0.714 | 3.83E-12 | 0.464 |  | 20:12 |  |
| *C8orf44* | HC/HF | 0.509 | 3.19E-07 | 0.410 | 0.503 | 03:19 | **0.012** |
|  | HF/HC | 0.730 | 1.18E-12 | 0.353 |  | 04:46 |  |
| *CBX4* | HC/HF | 0.275 | 1.17E-03 | 0.385 | 0.236 | 14:37 | **0.016** |
|  | HF/HC | 0.471 | 1.56E-06 | 0.252 |  | 17:16 |  |
| *CCNG2* | HC/HF | 0.496 | 5.68E-07 | 0.245 | 0.633 | 05:03 | **0.000** |
|  | HF/HC | 0.546 | 6.37E-08 | 0.284 |  | 11:11 |  |
| *CDC20* | HC/HF | 0.356 | 9.56E-05 | 0.232 | 0.809 | 15:40 | **0.000** |
|  | HF/HC | 0.278 | 1.08E-03 | 0.207 |  | 22:37 |  |
| *CEP76* | HC/HF | 0.374 | 5.36E-05 | 0.231 | 0.701 | 16:06 | **0.040** |
|  | HF/HC | 0.664 | 1.14E-10 | 0.258 |  | 18:21 |  |
| *COL25A1* | HC/HF | 0.344 | 1.44E-04 | 0.308 | 0.631 | 03:21 | **0.000** |
|  | HF/HC | 0.193 | 1.11E-02 | 0.251 |  | 13:10 |  |
| *CPEB2* | HC/HF | 0.431 | 7.07E-06 | 0.236 | 0.663 | 15:31 | **0.007** |
|  | HF/HC | 0.763 | 7.46E-14 | 0.262 |  | 17:21 |  |
| *DCUN1D3* | HC/HF | 0.090 | 1.38E-01 | 0.290 | 0.328 | 14:41 | **0.000** |
|  | HF/HC | 0.521 | 1.92E-07 | 0.458 |  | 16:34 |  |
| *DDIT3* | HC/HF | 0.478 | 1.16E-06 | 0.370 | 0.412 | 14:47 | **0.014** |
|  | HF/HC | 0.642 | 4.19E-10 | 0.307 |  | 15:55 |  |
| *FBXO6* | HC/HF | 0.252 | 2.22E-03 | 0.203 | 0.998 | 13:31 | **0.000** |
|  | HF/HC | 0.152 | 3.15E-02 | 0.203 |  | 02:13 |  |
| *GTPBP4* | HC/HF | 0.624 | 1.19E-09 | 0.213 | **0.010** | 20:14 | **0.003** |
|  | HF/HC | 0.660 | 1.41E-10 | 0.374 |  | 16:27 |  |
| *HEYL* | HC/HF | 0.088 | 1.44E-01 | 0.248 | 0.466 | 15:04 | 0.138 |
|  | HF/HC | 0.410 | 1.53E-05 | 0.369 |  | 17:06 |  |
| *HMGB2* | HC/HF | 0.462 | 2.24E-06 | 0.206 | **0.008** | 09:59 | 0.232 |
|  | HF/HC | 0.644 | 3.80E-10 | 0.384 |  | 07:43 |  |
| *HMGCS1* | HC/HF | 0.312 | 3.85E-04 | 0.211 | 0.760 | 23:49 | **0.011** |
|  | HF/HC | 0.549 | 5.55E-08 | 0.234 |  | 19:34 |  |
| *HOMEZ* | HC/HF | 0.264 | 1.58E-03 | 0.307 | 0.681 | 00:58 | **0.000** |
|  | HF/HC | 0.283 | 9.13E-04 | 0.254 |  | 16:28 |  |
| *IL16* | HC/HF | 0.333 | 2.04E-04 | 0.208 | 0.779 | 04:24 | **0.004** |
|  | HF/HC | 0.622 | 1.31E-09 | 0.227 |  | 08:31 |  |
| *IRS1* | HC/HF | 0.523 | 1.75E-07 | 0.439 | **0.002** | 20:16 | **0.008** |
|  | HF/HC | 0.755 | 1.43E-13 | 0.723 |  | 19:34 |  |
| *IRS2* | HC/HF | 0.711 | 4.85E-12 | 0.559 | **0.040** | 06:34 | 0.570 |
|  | HF/HC | 0.928 | 1.11E-16 | 0.718 |  | 07:03 |  |
| *KIAA0232* | HC/HF | 0.560 | 3.27E-08 | 0.210 | **0.000** | 17:10 | **0.013** |
|  | HF/HC | 0.854 | 1.11E-16 | 0.590 |  | 15:25 |  |
| *KLF11* | HC/HF | 0.211 | 6.81E-03 | 0.326 | 0.565 | 13:13 | 0.089 |
|  | HF/HC | 0.651 | 2.47E-10 | 0.398 |  | 11:50 |  |
| *KLHL31* | HC/HF | 0.163 | 2.40E-02 | 0.295 | 0.074 | 22:20 | 0.327 |
|  | HF/HC | 0.760 | 9.99E-14 | 0.537 |  | 19:56 |  |
| *L3HYPDH* | HC/HF | 0.194 | 1.09E-02 | 0.236 | 0.709 | 01:09 | **0.001** |
|  | HF/HC | 0.587 | 8.69E-09 | 0.206 |  | 19:53 |  |
| *LINC00672* | HC/HF | 0.426 | 8.51E-06 | 0.451 | 0.104 | 14:08 | **0.013** |
|  | HF/HC | 0.543 | 7.35E-08 | 0.297 |  | 12:53 |  |
| *LINC00842* | HC/HF | 0.123 | 6.33E-02 | 0.285 | 0.346 | 14:53 | 0.081 |
|  | HF/HC | 0.358 | 9.17E-05 | 0.474 |  | 17:12 |  |
| *LINC00989* | HC/HF | 0.344 | 1.44E-04 | 0.218 | 0.395 | 11:06 | **0.000** |
|  | HF/HC | 0.410 | 1.52E-05 | 0.307 |  | 04:31 |  |
| *LINC01128* | HC/HF | 0.458 | 2.64E-06 | 0.312 | 0.728 | 23:39 | **0.000** |
|  | HF/HC | 0.359 | 8.87E-05 | 0.274 |  | 16:40 |  |
| *LOC101928370* | HC/HF | 0.091 | 1.36E-01 | 0.209 | **0.000** | 02:21 | 0.974 |
|  | HF/HC | 0.657 | 1.75E-10 | 0.645 |  | 02:20 |  |
| *LOC101929018* | HC/HF | 0.410 | 1.53E-05 | 0.415 | 0.609 | 03:41 | **0.000** |
|  | HF/HC | 0.125 | 6.10E-02 | 0.330 |  | 13:45 |  |
| *LOC102723973* | HC/HF | 0.377 | 4.79E-05 | 0.330 | **0.013** | 09:41 | 0.163 |
|  | HF/HC | 0.738 | 6.06E-13 | 0.581 |  | 07:09 |  |
| *LOC102724679* | HC/HF | 0.261 | 1.77E-03 | 0.327 | 0.470 | 13:08 | **0.006** |
|  | HF/HC | 0.252 | 2.22E-03 | 0.235 |  | 15:58 |  |
| *LPIN1* | HC/HF | 0.505 | 3.88E-07 | 0.218 | 0.941 | 03:34 | **0.004** |
|  | HF/HC | 0.168 | 2.08E-02 | 0.212 |  | 01:14 |  |
| *MACC1* | HC/HF | 0.071 | 2.12E-01 | 0.327 | 0.650 | 02:12 | **0.000** |
|  | HF/HC | 0.306 | 4.68E-04 | 0.243 |  | 18:01 |  |
| *MIR3192* | HC/HF | 0.399 | 2.27E-05 | 0.293 | 0.312 | 13:09 | **0.017** |
|  | HF/HC | 0.467 | 1.85E-06 | 0.210 |  | 10:04 |  |
| *MIR3671* | HC/HF | 0.185 | 1.36E-02 | 0.287 | 0.516 | 03:46 | 0.059 |
|  | HF/HC | 0.525 | 1.65E-07 | 0.376 |  | 06:36 |  |
| *MIR3926-2* | HC/HF | 0.254 | 2.10E-03 | 0.277 | 0.559 | 13:54 | **0.000** |
|  | HF/HC | 0.217 | 5.82E-03 | 0.220 |  | 02:14 |  |
| *MIR518B* | HC/HF | 0.256 | 1.99E-03 | 0.224 | 0.740 | 03:35 | **0.000** |
|  | HF/HC | 0.316 | 3.39E-04 | 0.255 |  | 14:45 |  |
| *MSANTD4* | HC/HF | 0.329 | 2.27E-04 | 0.250 | 0.927 | 16:17 | **0.038** |
|  | HF/HC | 0.593 | 6.47E-09 | 0.242 |  | 19:20 |  |
| *NCAM1* | HC/HF | 0.829 | 1.11E-16 | 0.244 | 0.445 | 06:48 | **0.045** |
|  | HF/HC | 0.494 | 6.23E-07 | 0.207 |  | 04:50 |  |
| *NOP2* | HC/HF | 0.591 | 7.05E-09 | 0.533 | 0.206 | 16:10 | **0.000** |
|  | HF/HC | 0.673 | 6.38E-11 | 0.402 |  | 20:16 |  |
| *NR1D1* | HC/HF | 0.724 | 1.84E-12 | 0.981 | **0.008** | 07:18 | 0.270 |
|  | HF/HC | 0.876 | 1.11E-16 | 1.325 |  | 08:26 |  |
| *OR1D4* | HC/HF | 0.260 | 1.79E-03 | 0.261 | 0.723 | 01:26 | **0.000** |
|  | HF/HC | 0.219 | 5.61E-03 | 0.225 |  | 14:18 |  |
| *PCDH18* | HC/HF | 0.468 | 1.78E-06 | 0.421 | 0.202 | 23:37 | 0.293 |
|  | HF/HC | 0.717 | 3.06E-12 | 0.585 |  | 22:31 |  |
| *PCK1* | HC/HF | 0.386 | 3.62E-05 | 0.678 | **0.006** | 20:49 | 0.613 |
|  | HF/HC | 0.808 | 8.88E-16 | 1.120 |  | 19:53 |  |
| *PDK4* | HC/HF | 0.489 | 7.52E-07 | 0.955 | 0.146 | 03:44 | **0.000** |
|  | HF/HC | 0.875 | 1.11E-16 | 0.716 |  | 07:39 |  |
| *PHF12* | HC/HF | 0.401 | 2.12E-05 | 0.250 | 0.508 | 16:18 | **0.007** |
|  | HF/HC | 0.538 | 8.92E-08 | 0.304 |  | 14:33 |  |
| *PHF23* | HC/HF | 0.438 | 5.64E-06 | 0.349 | 0.752 | 15:41 | **0.002** |
|  | HF/HC | 0.623 | 1.30E-09 | 0.320 |  | 19:05 |  |
| *PIGC* | HC/HF | 0.211 | 6.89E-03 | 0.264 | 0.541 | 13:29 | **0.000** |
|  | HF/HC | 0.345 | 1.40E-04 | 0.202 |  | 04:05 |  |
| *PIN4* | HC/HF | 0.275 | 1.16E-03 | 0.325 | 0.344 | 03:06 | **0.000** |
|  | HF/HC | 0.216 | 6.03E-03 | 0.219 |  | 14:39 |  |
| *PPP1R15B* | HC/HF | 0.459 | 2.53E-06 | 0.243 | 0.644 | 15:20 | **0.033** |
|  | HF/HC | 0.826 | 1.11E-16 | 0.268 |  | 16:20 |  |
| *PRR4* | HC/HF | 0.116 | 7.42E-02 | 0.442 | 0.343 | 13:54 | **0.002** |
|  | HF/HC | 0.638 | 5.30E-10 | 0.642 |  | 16:30 |  |
| *REST* | HC/HF | 0.498 | 5.11E-07 | 0.214 | 0.210 | 15:58 | **0.007** |
|  | HF/HC | 0.808 | 8.88E-16 | 0.280 |  | 17:53 |  |
| *RFTN2* | HC/HF | 0.719 | 2.60E-12 | 0.403 | 0.068 | 07:56 | **0.001** |
|  | HF/HC | 0.701 | 9.51E-12 | 0.570 |  | 04:34 |  |
| *RPGR* | HC/HF | 0.419 | 1.13E-05 | 0.265 | 0.446 | 11:52 | 0.121 |
|  | HF/HC | 0.605 | 3.43E-09 | 0.333 |  | 09:52 |  |
| *RRS1* | HC/HF | 0.406 | 1.78E-05 | 0.476 | 0.334 | 15:47 | **0.002** |
|  | HF/HC | 0.680 | 4.13E-11 | 0.367 |  | 19:36 |  |
| *SFXN1* | HC/HF | 0.328 | 2.38E-04 | 0.317 | 0.640 | 14:57 | **0.008** |
|  | HF/HC | 0.506 | 3.63E-07 | 0.271 |  | 17:43 |  |
| *SLC10A6* | HC/HF | 0.308 | 4.34E-04 | 0.317 | 0.756 | 03:46 | **0.013** |
|  | HF/HC | 0.614 | 2.06E-09 | 0.286 |  | 07:04 |  |
| *SLC25A15* | HC/HF | 0.315 | 3.52E-04 | 0.234 | 0.828 | 00:03 | **0.000** |
|  | HF/HC | 0.469 | 1.67E-06 | 0.254 |  | 16:03 |  |
| *SNORD124* | HC/HF | 0.261 | 1.76E-03 | 0.215 | 0.985 | 15:26 | **0.000** |
|  | HF/HC | 0.194 | 1.07E-02 | 0.213 |  | 00:43 |  |
| *SNORD14E* | HC/HF | 0.057 | 2.93E-01 | 0.238 | 0.670 | 15:04 | **0.007** |
|  | HF/HC | 0.342 | 1.54E-04 | 0.315 |  | 21:16 |  |
| *SOCS4* | HC/HF | 0.318 | 3.20E-04 | 0.203 | 0.826 | 16:23 | 0.060 |
|  | HF/HC | 0.623 | 1.26E-09 | 0.218 |  | 19:03 |  |
| *SOS2* | HC/HF | 0.341 | 1.58E-04 | 0.240 | 0.753 | 14:14 | **0.006** |
|  | HF/HC | 0.471 | 1.56E-06 | 0.218 |  | 12:42 |  |
| *TFPI2* | HC/HF | 0.244 | 2.79E-03 | 0.570 | 0.798 | 00:56 | **0.006** |
|  | HF/HC | 0.696 | 1.37E-11 | 0.525 |  | 21:23 |  |
| *THRSP* | HC/HF | 0.385 | 3.64E-05 | 0.386 | 0.146 | 17:40 | 0.103 |
|  | HF/HC | 0.725 | 1.66E-12 | 0.556 |  | 20:12 |  |
| *TP53RK* | HC/HF | 0.382 | 4.08E-05 | 0.387 | 0.242 | 15:15 | **0.029** |
|  | HF/HC | 0.493 | 6.31E-07 | 0.265 |  | 17:40 |  |
| *UBA6-AS1* | HC/HF | 0.398 | 2.35E-05 | 0.320 | 0.851 | 03:34 | **0.013** |
|  | HF/HC | 0.278 | 1.08E-03 | 0.341 |  | 02:04 |  |
| *USP38* | HC/HF | 0.379 | 4.58E-05 | 0.221 | 0.174 | 15:42 | 0.367 |
|  | HF/HC | 0.655 | 1.96E-10 | 0.322 |  | 16:18 |  |
| *WASH3P* | HC/HF | 0.147 | 3.53E-02 | 0.203 | 0.705 | 04:29 | **0.000** |
|  | HF/HC | 0.228 | 4.36E-03 | 0.260 |  | 17:04 |  |
| *ZDHHC14* | HC/HF | 0.543 | 7.38E-08 | 0.409 | **0.045** | 04:49 | 0.888 |
|  | HF/HC | 0.431 | 7.26E-06 | 0.210 |  | 05:00 |  |
| *ZNF503* | HC/HF | 0.428 | 7.91E-06 | 0.277 | 0.361 | 14:37 | **0.019** |
|  | HF/HC | 0.431 | 7.14E-06 | 0.373 |  | 16:08 |  |
| *ZNF674-AS1* | HC/HF | 0.384 | 3.87E-05 | 0.325 | 0.342 | 14:40 | **0.014** |
|  | HF/HC | 0.332 | 2.12E-04 | 0.242 |  | 13:20 |  |
| *ZNF791* | HC/HF | 0.333 | 2.01E-04 | 0.225 | 0.942 | 14:05 | **0.007** |
|  | HF/HC | 0.479 | 1.14E-06 | 0.230 |  | 12:30 |  |
| *ZNF844* | HC/HF | 0.413 | 1.37E-05 | 0.232 | 0.519 | 04:57 | **0.029** |
|  | HF/HC | 0.428 | 7.91E-06 | 0.296 |  | 09:02 |  |

Gene expression was analysed on the microarray dataset (n=15 per time point) and the oscillation fit was performed using a sinus model (cosinor method). Significant oscillation was considered when amplitude ≥ 0.2 and p<0.05. Amplitude is the distance between the peak and the mean value of a wave; Acrophase refers to the time at which the peak occurs ((h): hours). The first P column indicates significant diurnal oscillation for each group. The P (Amp) and P (Acr) columns reflect the statistical differences between groups for each circadian parameter by cosinor method (amplitude and acrophase, respectively). * p-values without correction for multiple testing are shown; no significant differences were found after FDR correction. HC, high carbohydrate; HF, High fat.

**Table S4.** **Oscillation parameters of transcripts in SAT and PBMC and biomarkers in saliva and serum.**

| **Parameter** | **Diet** | **Cosinor fitting** | | **Amplitude** | **P (Amp)*** | **Acrophase/nadir (h)** | **P (Acr)*** |
| --- | --- | --- | --- | --- | --- | --- | --- |
|  |  | **r^2^** | **P*** |  |  |  |  |
| Subcutaneous adipose tissue | | | | | | | |
| *BMAL1/ARNTL* | HC/HF | 0.39 | <0.001 | 0.38 | 0.973 | 20:07 | 0.496 |
|  | HF/HC | 0.44 | <0.001 | 0.38 |  | 21:44 |  |
| *CLOCK* | HC/HF | 0.30 | 0.001 | 0.16 | 0.959 | 23:05 | 0.321 |
|  | HF/HC | 0.33 | <0.001 | 0.16 |  | 20:37 |  |
| *CRY1* | HC/HF | 0.41 | <0.001 | 0.35 | 0.584 | 16:09 | 0.486 |
|  | HF/HC | 0.72 | <0.001 | 0.44 |  | 15:42 |  |
| *CRY2* | HC/HF | 0.28 | <0.001 | 0.28 | 0.478 | 11:26 | 0.377 |
|  | HF/HC | 0.86 | <0.001 | 0.36 |  | 12:18 |  |
| *DBP* | HC/HF | 0.92 | <0.001 | 0.61 | 0.554 | 7:58 | 0.734 |
|  | HF/HC | 0.64 | <0.001 | 0.58 |  | 8:13 |  |
| *NR1D1* | HC/HF | 0.72 | <0.001 | 0.98 | **0.008** | 7:19 | 0.270 |
|  | HF/HC | 0.87 | <0.001 | 1.33 |  | 8:26 |  |
| *NR1D2* | HC/HF | 0.72 | <0.001 | 0.53 | 0.341 | 7:36 | 0.682 |
|  | HF/HC | 0.91 | <0.001 | 0.59 |  | 8:01 |  |
| *PER1* | HC/HF | 0.78 | <0.001 | 0.98 | 0.339 | 8:58 | 0.867 |
|  | HF/HC | 0.85 | <0.001 | 1.09 |  | 8:48 |  |
| *PER2* | HC/HF | 0.47 | <0.001 | 0.38 | **0.026** | 10:42 | 0.836 |
|  | HF/HC | 0.76 | <0.001 | 0.67 |  | 11:39 |  |
| *PER3* | HC/HF | 0.63 | <0.001 | 0.48 | 0.220 | 9:16 | 0.484 |
|  | HF/HC | 0.76 | <0.001 | 0.60 |  | 10:08 |  |
| *TEF* | HC/HF | 0.38 | <0.001 | 0.23 | **0.033** | 7:59 | 0.176 |
|  | HF/HC | 0.75 | <0.001 | 0.39 |  | 10:27 |  |
| Peripheral blood mononuclear cells | | | | | | | |
| *PER1* | HC/HF | 0.27 | 0.002 | 0.46 | 0.670 | 9:49 | **0.008** |
|  | HF/HC | 0.27 | 0.002 | 0.56 |  | 4:06 |  |
| *IRS1* | HC/HF | 0.22 | 0.006 | 0.86 | 0.602 | 11:59 | 0.637 |
|  | HF/HC | 0.14 | 0.047 | 0.62 |  | 12:44 |  |
| *PDK4* | HC/HF | 0.43 | <0.001 | 1.14 | **0.014** | 3:52 | **0.001** |
|  | HF/HC | 0.58 | <0.001 | 0.54 |  | 8:31 |  |
| Salivary cortisol and serum IL-6 | | | | | | | |
| Cortisol | HC/HF | 0.51 | <0.001 | 0.27 | 0.835 | 7:11 | 0.490 |
|  | HF/HC | 0.58 | <0.001 | 0.26 |  | 6:46 |  |
| IL-6 | HC/HF | 0.23 | <0.001 | 1.46 | 0.949 | 17:38 | 0.321 |
|  | HF/HC | 0.18 | <0.001 | 1.43 |  | 19:37 |  |

Oscillation parameters were analysed in SAT (n=15) and peripheral blood mononuclear cell transcripts (n=15) as well as in salivary cortisol and serum IL-6 (n=15). For cortisol – nadir times because a nadir (time of the curve minimum) is a more robust marker of phase of central clock compared with cortisol acrophase. The oscillation fit was performed using a sinus model (cosinor method). Significant oscillation was considered when amplitude ≥ 0.2 and p<0.05. Amplitude is the distance between the peak and the mean value of a wave; acrophase refers to the time at which the peak occurs ((h): hours). The first P column indicates significant diurnal oscillation for each group. The P (Amp) and P (Acr) columns reflect the statistical differences between groups for each circadian parameter by cosinor method (amplitude and acrophase, respectively). * p-values without correction for multiple testing are shown; no significant differences were found after the false discovery rate correction. HC, high-carb; HF, high-fat.

**Table S4. Primers used for real-time PCR.**

| **Gene name** | **Gene symbol** | **Gene ID** | **Primer sequence** | |
| --- | --- | --- | --- | --- |
|  |  |  | **forward** | **reverse** |
| **Clock genes** |  |  |  |  |
| Aryl hydrocarbon receptor nuclear translocator-like | *BMAL1/ARNTL* | 406 | CATTAAGAGGTGCCACCAATCC | CAAAAATCCATCTGCTGCCC |
| Nuclear receptor subfamily 1, group D, member 1 | *NR1D1* | 9572 | TGACCTTTCTCAGCATGACCAA | CAAAGCGCACCATCAGCAC |
| Period circadian clock 1 | *PER1* | 5187 | ATTCCGCCTAACCCCGTATGT | CCGCGTAGTGAAAATCCTCTTG |
| **Glucose and lipid metabolism genes** | | | | |
| Insulin receptor substrate 1 | *IRS1* | 3667 | CGAAAGAGAACTCACTCGGCA | TAGGCAGGCATCATCTCTGTGT |
| Insulin receptor substrate 2 | *IRS2* | 8660 | ACGAGAGCGAGAAAAAGTGGC | ATCAGGTACTTGTGCTTGGCG |
| Phosphoenolpyruvate carboxykinase 1 | *PCK1* | 5105 | AAGTATGACAACTGCTGGTTGGC | ATAACCGTCTTGCTTTCGATCCT |
| Pyruvate dehydrogenase kinase 4 | *PDK4* | 5166 | CCCTGAGAATTATTGACCGCCT | AAGCCGTAACCAAAACCAGCC |
| **Inflammatory genes and adipokines** | | | | |
| C-C motif chemokine ligand 5 | *CCL5* | 6352 | CGTGCCCACATCAAGGAG | GGACAAGAGCAAGCAGAAAC |
| Interleukin 1 beta | *IL1B* | 3553 | GCAATGAGGATGACTTGTTCTTTG | CAGAGGTCCAGGTCCTGGAA |
| Protein kinase C beta | *PRKCB* | 5579 | ACCGCCTGTACTTTGTGATGGAGT | CCGATGGCAATTTCTGCAGCGTAA |
| NLR family pyrin domain containing 3 | *NLRP3* | 114548 | AAAGAGATGAGCCGAAGTGGG | TCAATGCTGTCTTCCTGGCA |
| Toll like receptor 2 | *TLR2* | 7097 | AGCACTGGACAATGCCACATAT | CATTGCGGTCACAAGACAGAGA |
| **Housekeeping genes** | | | | |
| Beta-2-microglobulin | *B2M* | 567 | CTATCCAGCGTACTCCAAAG | AAACCCAGACACATAGCAAT |
| Glucuronidase, beta | *GUSB* | 2990 | CTCATTTGGAATTTTGCCGATT | CCGAGTGAAGATCCCCTTTTTA |

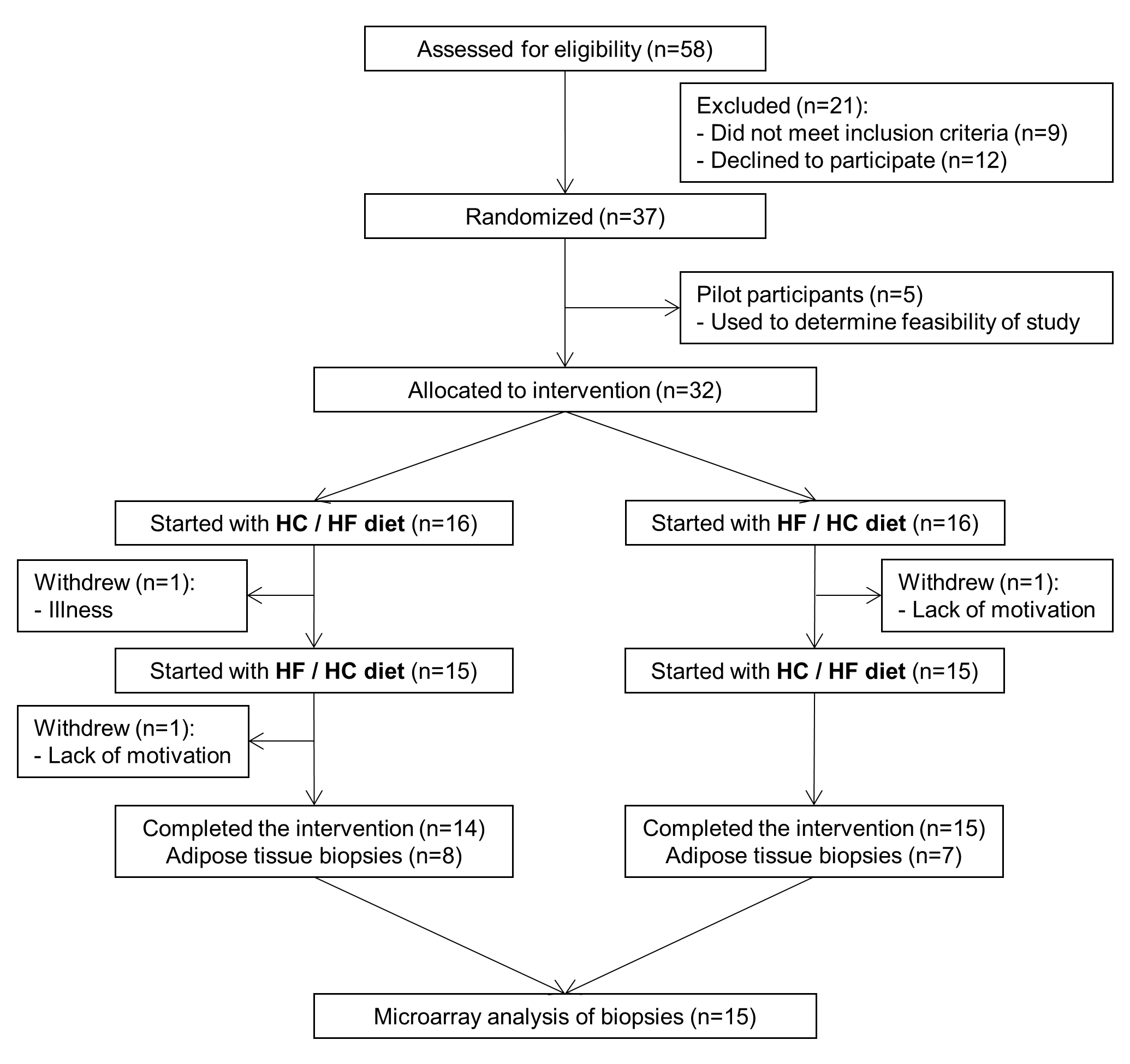

**Figure S1. Study flow chart.**

A total of 58 subjects were assessed for eligibility, 37 of whom were considered eligible. 5 men were used in a pilot project to determine feasibility of study. 3 subjects withdrew from the study, thus totally 29 subjects completed the study. Adipose tissue biopsies were collected in 15 subjects for transcriptome analysis. HC/HF diet, isocaloric high-carbohydrates meals from 6:00 to 13:30 and isocaloric high-fat meals between 16.30 and 22:00; HF/HC diet, reversed order of meal sequence.

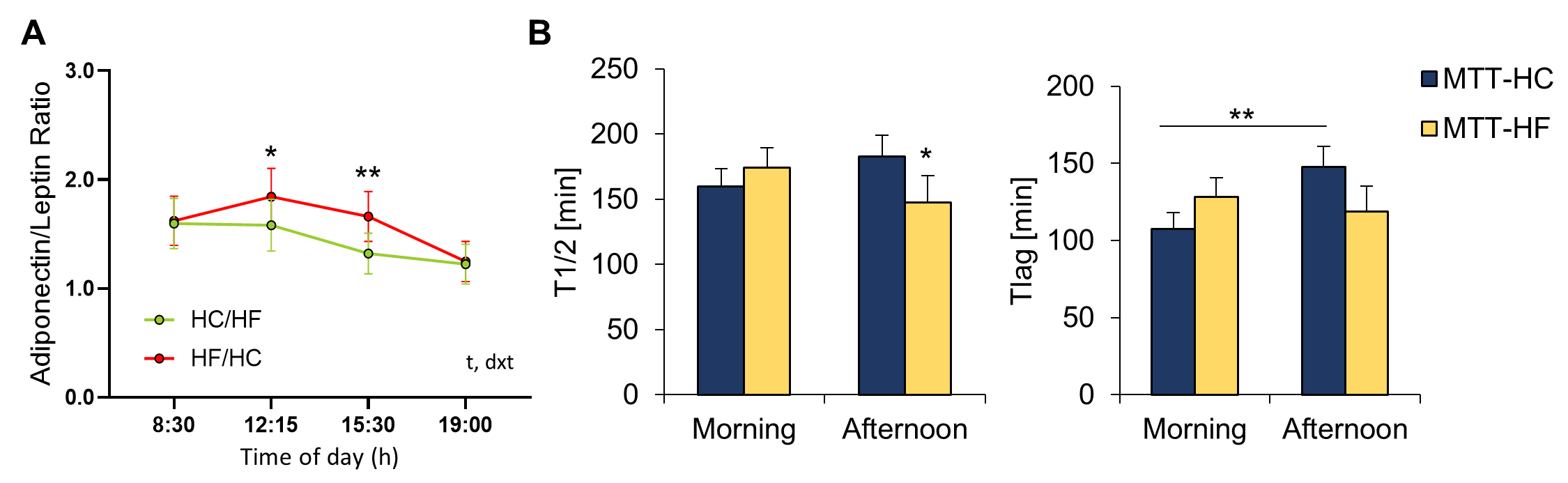

**Figure S2. Adiponectin/leptin ratio and gastric emptying rates after meal tolerance tests (n=15).**

(A) Serum levels throughout the day of the adiponectin/leptin ratio after HC/HF (green) and HF/HC (red) diets. Data are expressed as mean ± SEM for each time point. p-values were calculated by two-way repeated-measures ANOVA (t, by time; dxt, by diet*time interaction). * p<0.05, *** p<0.001 indicate significant difference between HC/HF diet vs. HF/HC diet at the same time of the day by paired t-test. HC, high carbohydrate; HF, high fat.

(B) At the end of each intervention period, two meal tolerance tests, carbohydrate-rich (MTT-HC, blue) or fat-rich (MTT-HF, yellow), were performed at 09.00 am and 03.40 pm during the investigation day. T1/2 - half gastric emptying time; Tlag - time of fastest gastric emptying (n=29). * p<0.05; ** p<0.01 by paired t-test. Data are means ± SEM.

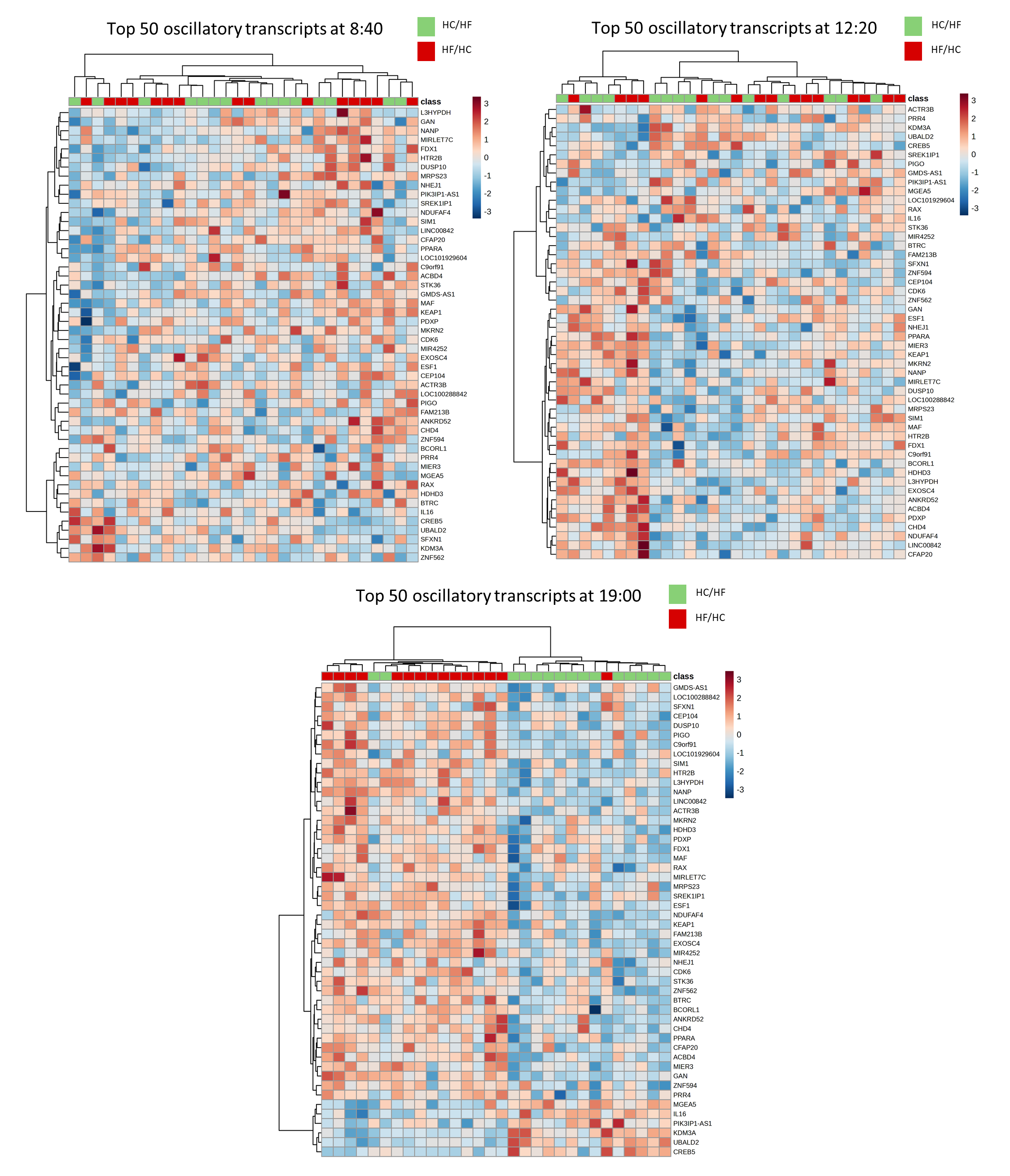

**Figure S3. Meal composition change the transcriptome profile in adipose tissue at 19:00 time point (n=15).**

Heatmap representation of the top 50 oscillatory transcripts after each diet at the 8:40, 12:20 and 19:00 time points.

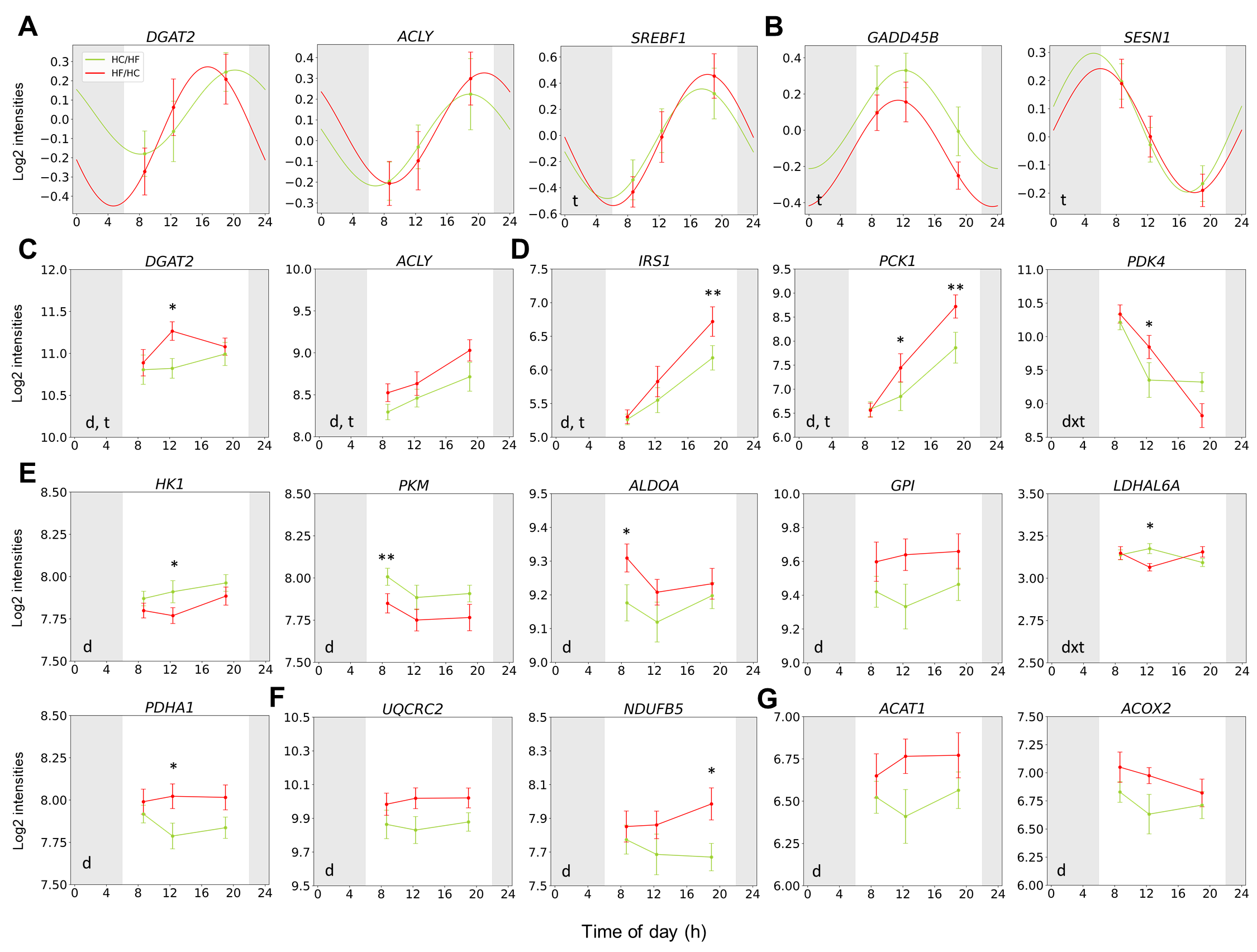

**Figure S4.** **Oscillating and non-oscillating genes involved in lipid and glucose metabolism in SAT (n=15)**.

(A) Diurnal oscillations of genes involved in lipogenesis after HC/HF (green) and HF/HC (red) diets.

(B) Diurnal oscillations of genes involved in lipid mobilization after HC/HF (green) and HF/HC (red) diets.

(C) Diurnal expression patterns of *DGAT2* and *ACLY* after HC/HF (green) and HF/HC (red) diets.

(D) Diurnal expression patterns of oscillating glucose and lipid metabolism genes after HC/HF (green) and HF/HC (red) diets. These graphs are shown additionally to oscillation graphs only for genes with significant diet or diet x time effects by two-way repeated-measures ANOVA.

(E) Diurnal expression patterns of genes involved in glycolysis and glucose utilization after HC/HF (green) and HF/HC (red) diets.

(F) Diurnal expression patterns of genes involved in mitochondrial electron transport and oxidative phosphorylation after HC/HF (green) and HF/HC (red) diets.

(G) Diurnal expression patterns of genes involved in lipogenesis and lipid oxidation after HC/HF (green) and HF/HC (red) diets.

Data are expressed as mean ± SEM for each time point. For oscillation transcripts, the oscillation fit was performed using a sinus model (cosinor method). Significant oscillation was considered when amplitude ≥ 0.2 and p<0.05. Significant changes of the oscillation acrophase (p) and/or amplitude (a) are shown. The full data analysis is shown in Supplementary Dataset 2. p-values were calculated by two-way repeated-measures ANOVA (d, by diet; t, by time). * p<0.05, ** p<0.01 indicate significant difference between HC/HF diet vs. HF/HC diet at the same time of the day by paired t-test.

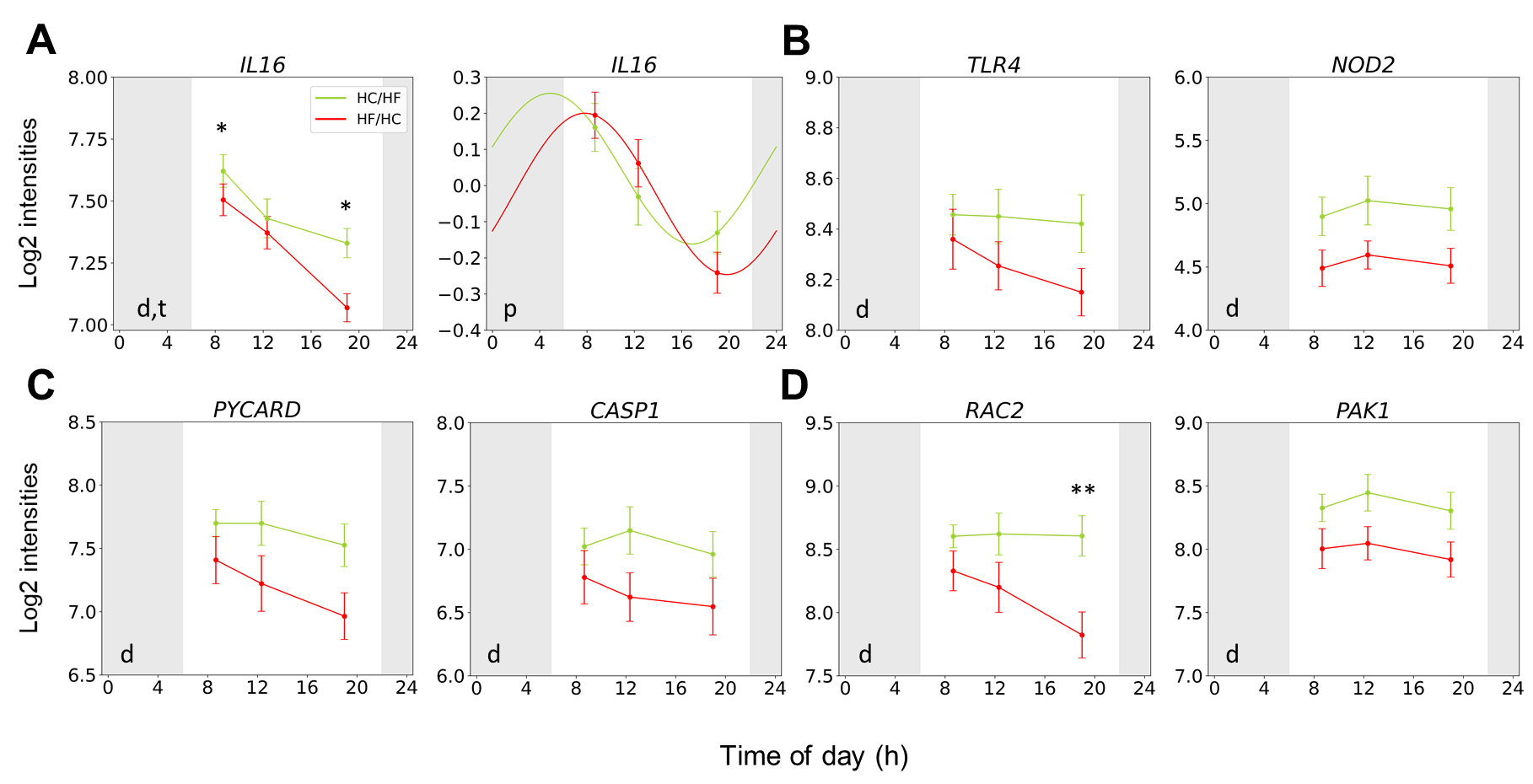

**Figure S5. Oscillating and non-oscillating genes involved in inflammatory response in SAT (n=15).**

(A) Diurnal expression patterns and diurnal oscillation of *IL16* gene involved in inflammatory and chemotactic signalling after HC/HF (green) and HF/HC (red) diets.

(B) Diurnal expression patterns of genes involved in NOD-like receptor signalling and Toll-like receptor signalling after HC/HF (green) and HF/HC (red) diets.

(C) Diurnal expression patterns of genes involved in NLRP3 inflammasome after HC/HF (green) and HF/HC (red) diets.

(D) Diurnal expression patterns of genes involved in reactive oxygen species production and macrophage activation after HC/HF (green) and HF/HC (red) diets.

Data are expressed as mean ± SEM for each time point. Significant oscillation was considered when amplitude ≥ 0.2 and p<0.05. Significant changes of the oscillation acrophase (p) and/or amplitude (a) are shown. The full data analysis is shown in Supplementary Dataset 2. p-values were calculated by two-way repeated-measures ANOVA (d, by diet; t, by time). * p<0.05, ** p<0.01 indicate significant difference between HC/HF diet vs. HF/HC diet at the same time of the day by paired t-test.

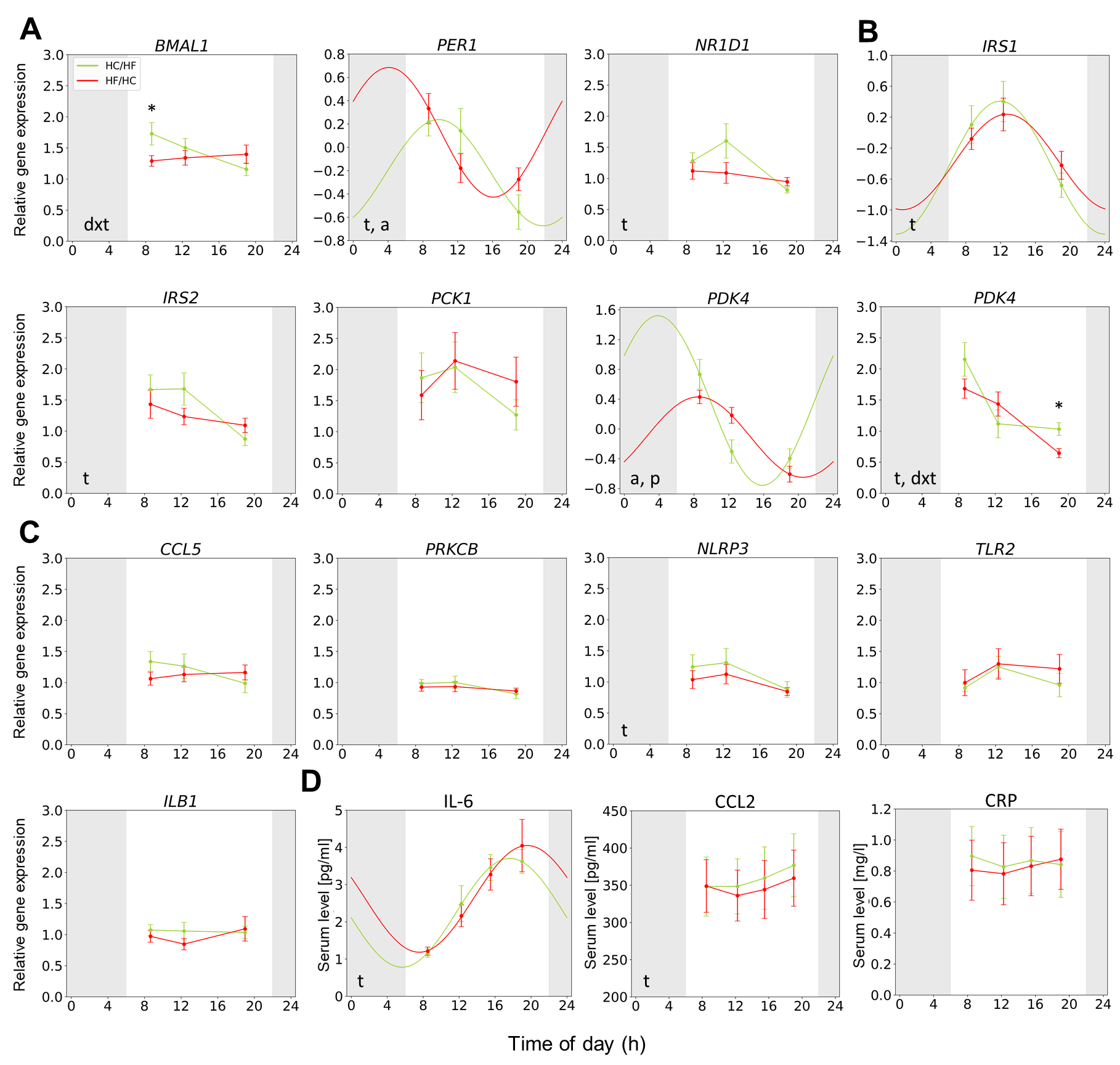

**Figure S6. Diet composition influenced diurnal oscillations and expression levels of glucose and lipid metabolism genes in PBMCs (n=15)**.

(A) Diurnal expression patterns and oscillations of clock genes after HC/HF (green) and HF/HC (red) diets.

(B) Diurnal expression patterns and oscillations of glucose and lipid metabolism genes and diurnal expression patterns of *PDK4* after HC/HF (green) and HF/HC (red) diets. These graphs are shown additionally to oscillation graphs only for genes with significant diet or diet x time effects by two-way repeated-measures ANOVA.

(C) Diurnal expression patterns and oscillations of genes involved in inflammatory response after HC/HF (green) and HF/HC (red) diets.

(D) Diurnal serum patterns and oscillations of inflammatory response parameters after HC/HF (green) and HF/HC (red) diets.

Data are expressed as mean ± SEM for each time point. For oscillation transcripts, the oscillation fit was performed using a sinus model (cosinor method). Significant oscillation was considered when amplitude ≥ 0.2 and p<0.05. Significant changes of the oscillation acrophase (a) and/or amplitude (p) are shown. The full data analysis is shown in Table S3. For non-oscillating transcripts, p-values were calculated by two-way repeated-measures ANOVA (d, by diet; t, by time; dxt, by diet*time interaction). * p<0.05, *** p<0.001 indicate significant difference between HC/HF diet vs. HF/HC diet at the same time of the day by paired t-test.
